# Cardiometabolic health trajectories from birth to old age based on multi-decadal series of biochemistry and anthropometry

**DOI:** 10.64898/2026.04.01.26349266

**Authors:** Ville-Petteri Mäkinen, Mika Kähönen, Terho Lehtimäki, Nina Hutri, Tapani Rönnemaa, Jorma Viikari, Katja Pahkala, Suvi Rovio, Harri Niinikoski, Juha Mykkänen, Olli Raitakari, Mika Ala-Korpela

**Author notes:** Corresponding author: Dr Ville-Petteri Mäkinen, Systems Epidemiology, Research Unit of Population Health, Faculty of Medicine, University of Oulu and Biocenter Oulu Oulu, Finland.

## Abstract

**Background and aims:** Direct evidence to connect early life metabolism with cardiometabolic diseases in old age is limited due to the rarity of multi-decadal biochemical follow-up studies. To gain deeper insight into metabolic ageing, we conducted a longitudinal study that integrates serial data on clinical biomarkers, metabolomics and clinical events across the human life course.

**Methods:** Children born in 1962–1992 were included from four European cohorts. Time-series of clinical biomarkers and metabolomics data were available for 8,653 participants (ages 0-49 years, 142 molecular and four physiological variables). Comparable data for 13,795 UK Biobank participants at two visits (ages 40-79 years) were linked with retrospective and prospective records of diabetes and cardiovascular disease. Lifetime metabolic trajectories were reconstructed by unsupervised machine learning and local polynomial regression.

**Results:** A stable stratification in metabolic health emerged in children between ages 3 and 12 years and persisted to old age. We summarized this population pattern by assigning each participant into one of seven metabolic subgroups with characteristic biomarker trajectories. Two subgroups (MetDys TG+ and MetDys TG−) featured increased waist-height ratio from childhood, persistently higher C-reactive protein throughout life and rapidly increasing fasting insulin between 30 and 49 years of age. Both subgroups exhibited high risk for diabetes (HR > 13) and ischemic heart disease (HR > 2.5) when compared against the lowest risk subgroup (High HDL ApoB−).

**Conclusions:** This life-course analysis shows that metabolic dysfunction associated with excess weight gain begins in early childhood and is associated with cardiometabolic morbidity in later life.

## Introduction

Longitudinal cohort studies have established predictive associations between early life cardiometabolic biomarkers and disease risk later in life that present an opportunity for preventative interventions [1–4]. Simultaneously, molecular profiling of children and adults has revealed considerable diversity in cardiometabolic health indicators with potentially profound implications for the timing and nature of these interventions. For example, a multi-omics study identified a high-risk subgroup of children with higher body mass, inflammation and dysfunction across multiple metabolic pathways that may provide new precision targets for early interventions [5] and large-scale analyses of adults have elucidated the connections between molecular diversity and cardiovascular risk stratification [6–10]. Aside from purely data-driven clustering analyses, genetic and biochemical studies have revealed molecular heterogeneity among individuals with obesity and diabetes [11], which has increased the interest in metabolically “healthy” versus “non-healthy” obesity subtypes as important refinements to the classical diagnostic categories [12].

The significance of metabolic diversity in children is particularly important now, given the current failure of societies to reverse obesity trends. More children with obesity today means that ever greater proportion of adults tomorrow are likely to be exposed to CVD drivers such as dyslipidaemia and diabetes [13,14]. Interestingly, recent advances in obesity medications may make population-wide weight reduction finally feasible [15] and the treatments can be started early in life [16]. Whether the new interventions should be translated into general CVD prevention remains an open question – a 50-year weight management trial to prevent CVD events would be necessary to measure effect sizes directly, but such evidence is out of reach [17]. In the absence of trial data, we postulate that careful analyses of the observational temporal trajectories that connect childhood metabolic stratification with diverging disease risk in adulthood will provide us with novel and useful insight into how cardiometabolic disease burden can be reduced.

Previous prospective studies have established predictive associations between early life factors and cardiovascular events later in life [1–4], but they offer limited insight into the temporal biomarker trajectories associated with the risk strata, and the results apply to only those individuals who were the first to be affected by CVD. Hence the picture of childhood risk factors in relation to the bulk of cardiovascular events at older ages remains incomplete. Indeed, we are not aware of any single large-scale cohort that would have comprehensive biochemical time-series data across several decades; modern techniques such as nuclear magnetic resonance (NMR) metabolomics [18] are relatively recent inventions. The gaps in temporal biochemical data on the path to clinical endpoints limit the evidence base for the lifetime risk factors of cardiovascular disease and hamper efforts to optimize screening for high-risk blood lipids and other biomarkers in children and adolescents [17].

To overcome the data limitations, we developed a novel integrative study design that leveraged multiple longitudinal human cohorts and enabled us to trace continuous metabolic health trajectories from birth to the eighth decade of life. The specific aims were 1) to stratify the study population according to longitudinal multivariable metabolic trajectories during the first half of life and 2) to identity those metabolic strata that manifest high rates of cardiometabolic diseases at older ages. We utilized a unique collection of five datasets that each included multiple time points with clinical biochemistry and metabolomics (total 22,448 participants). We also used a sophisticated neural network pipeline [19] to dissect multivariable temporal patterns of 146 metabolic measures, and to elucidate their associations with prevalent and incident diabetes and ischemic heart disease. This is the first systems epidemiological study that reveals the biochemical arc of life-long cardiometabolic health stratification and its clinical consequences from birth to old age.

## Materials and Methods

The study included participants from five datasets: The Avon Longitudinal Study of Parents and Children (ALSPAC) [20–23], The Special Turku Coronary Risk Factor Intervention Project (STRIP) [24,25], The Cardiovascular Risk in Young Finns Study (YFS) [26], The Northern Finland Birth Cohort 1966 (NFBC66) [27] and the UK Biobank project #324302 [27]. The temporal overlaps between the cohorts are illustrated in Figure 1A. Further details are available in Supplementary Table S1 and in Supplementary Figure S1.

**Figure 1:**
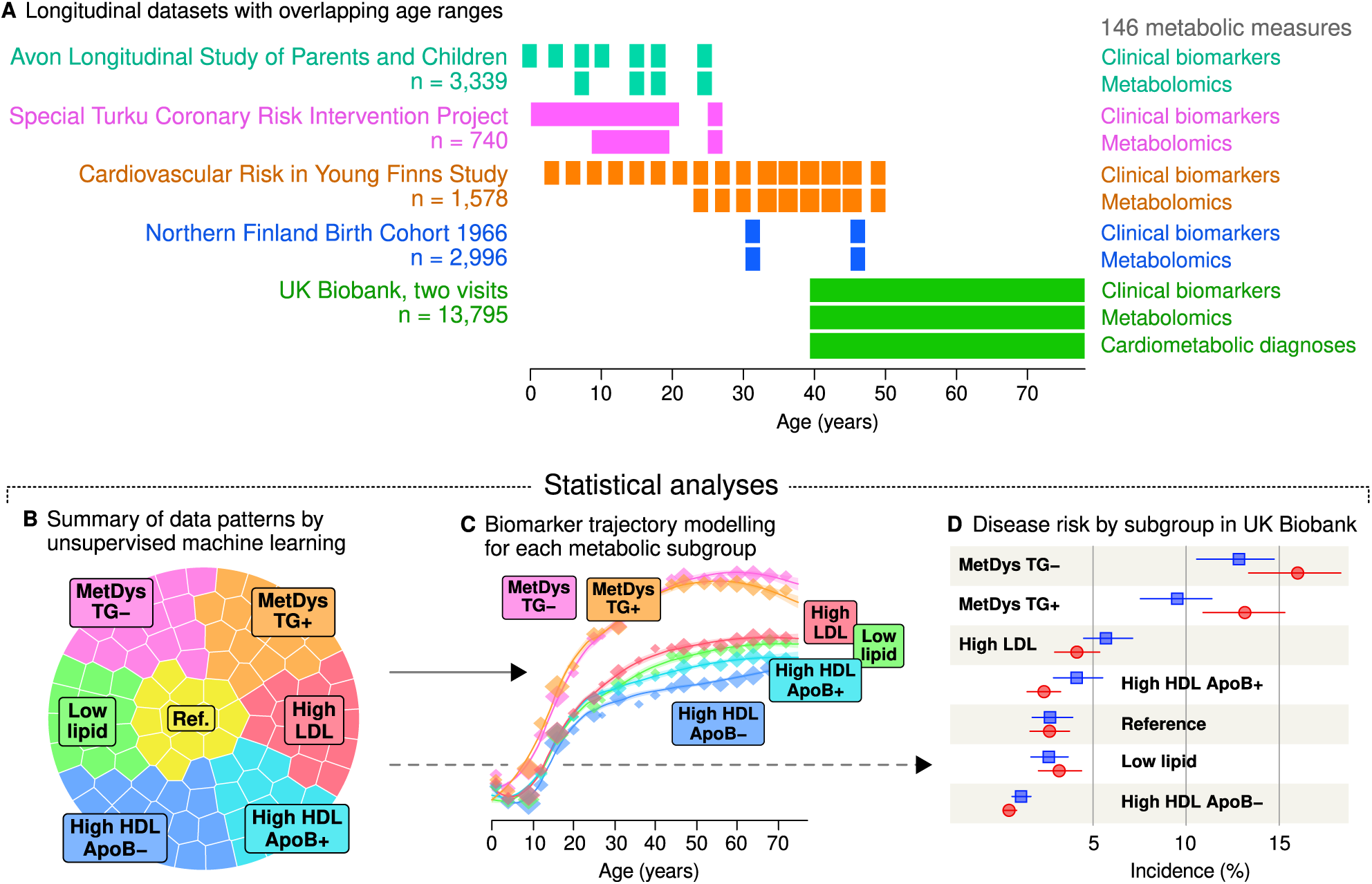
Schematic illustration of data coverage and statistical analyses. Longitudinal data were sourced from five European cohorts with overlapping age coverage (Plot A). The numbers of participants that were included in the study are listed, please see Supplementary Material for additional information on how data were filtered. In the first part of the study, all participants were stratified into seven subgroups based on their anthropometric and biochemical features (Plot B). Next, biomarkers with sufficient serial data available were chosen for lifetime trajectory reconstruction for each subgroup, respectively (Plot B). Lastly, the disease risk associated with each subgroup was assessed in the UK Biobank (Plot D). Abbreviations: apolipoprotein B (ApoB), high-density lipoprotein (HDL), low-density lipoprotein (LDL), metabolic dysfunction (MetDys), triglycerides (TG).

Height, weight, waist and hip circumference and calculated body-mass index (BMI) and waist-height ratio (WHER) were available from multiple time points. Standard clinical chemistry methods were used for 31 biomarkers, including triglycerides, high-density (HDL) and low-density lipoprotein (LDL) cholesterol, glucose, hemoglobin A1c, insulin, C-reactive protein and liver enzymes. NMR metabolomics data included 251 measures such as lipoprotein lipids, amino acids, ketones and creatinine [18]. Ratios of molecules were excluded to avoid redundancy when training multi-variate models. The final dataset for subgroup modelling contained 142 biochemical and four physiological measures (Supplementary Table S2).

### Data-driven multi-variable metabolic subgroups

Study participants were assigned to metabolic subgroups according to a self-organizing map (SOM). The SOM is an artificial neural network approach that is designed to facilitate the detection of multi-variable patterns in complex datasets and in this study we used the Numero R library that was specifically designed for epidemiological datasets [6,19]. The method produces a two-dimensional layout of the data where participants with similar metabolic profiles are close to each other on the map and thus can be assigned to the same subgroup by visually observable proximity.

Due to cohort design, attrition and changes in data collection over the years, data coverage varied across the decades. We selected a total of 8,653 individuals with the best coverage as the longitudinal dataset for trajectory modelling (Supplementary Figure S2). We chose i) variables that were available in at least two cohorts, ii) data points with body weight and biochemistry available at three or more ages, and iii) participants with data spanning at least eight years of follow-up.

Input data for the SOM were split into batches according to cohort, age group and sex, and each variable was log-transformed if skewed, centred to zero and scaled to unit variance. Collinearity among NMR-based measures was reduced by merging highly correlated variables into a single data column (Supplementary Figure S3). UK Biobank was not used for training the SOM, however, the metabolic data were pre-processed the same way such that the data could be projected onto the trained map. UK Biobank biochemical data were adjusted for lipid-lowering medication according to effect sizes from a metabolomics study of statins [28] and for anti-hypertensive medication according to Cochrane systematic reviews (Supplementary Table S3).

To incorporate the temporal dimension into the model, the input data matrix was reshaped as described in Supplementary Figure S4. The technical quality control methods and results for the SOM modelling are presented in the supplementary material, including Supplementary Figures S5 and S6. The subsequent analysis steps in the SOM framework followed the previously published protocol [19] and are described in Results.

### Subgroup trajectory modelling

Cubic splines and polynomial regression were used to investigate the temporal evolution of metabolic traits within a subgroup. Based on earlier findings [29,30], we set the spline knots at 2, 13, 21, 47 and 69 years of age, except for C-reactive protein, where we used 17, 23, 35, 47 and 69 years. Scale differences in biomarker values between cohorts were adjusted by first fitting the spline model to log-transformed data with indicators for cohort membership and then adjusting the original scales according to the indicator coefficients (referenced to YFS). Final subgroup trajectories were fitted to the scaled data using local polynomial regression [31]. Both clinical assays and NMR measures were available for triglycerides, cholesterol, LDL and HDL cholesterol, apolipoprotein B and glucose; for maximal trajectory coverage, missing clinical values were imputed from NMR data by linear regression where available.

All analyses were conducted using the R statistical environment version 4 (The R Foundation, Institute for Statistics and Mathematics, Wirtschaftsuniversität Wien, Vienna, Austria).

## Results

We defined seven regions based on SOM visualizations (with priority given to biomarkers of clinical significance) and assigned the participants located in each region to a separate subgroup. The centre of the map was chosen as the population “reference” or average category (Figure 2D,J,P,V), since these individuals were the most likely to have near-average values across all metabolic measures simultaneously. The reference subgroup was omitted from figures for better readability, but included in tables and other results.

**Figure 2:**
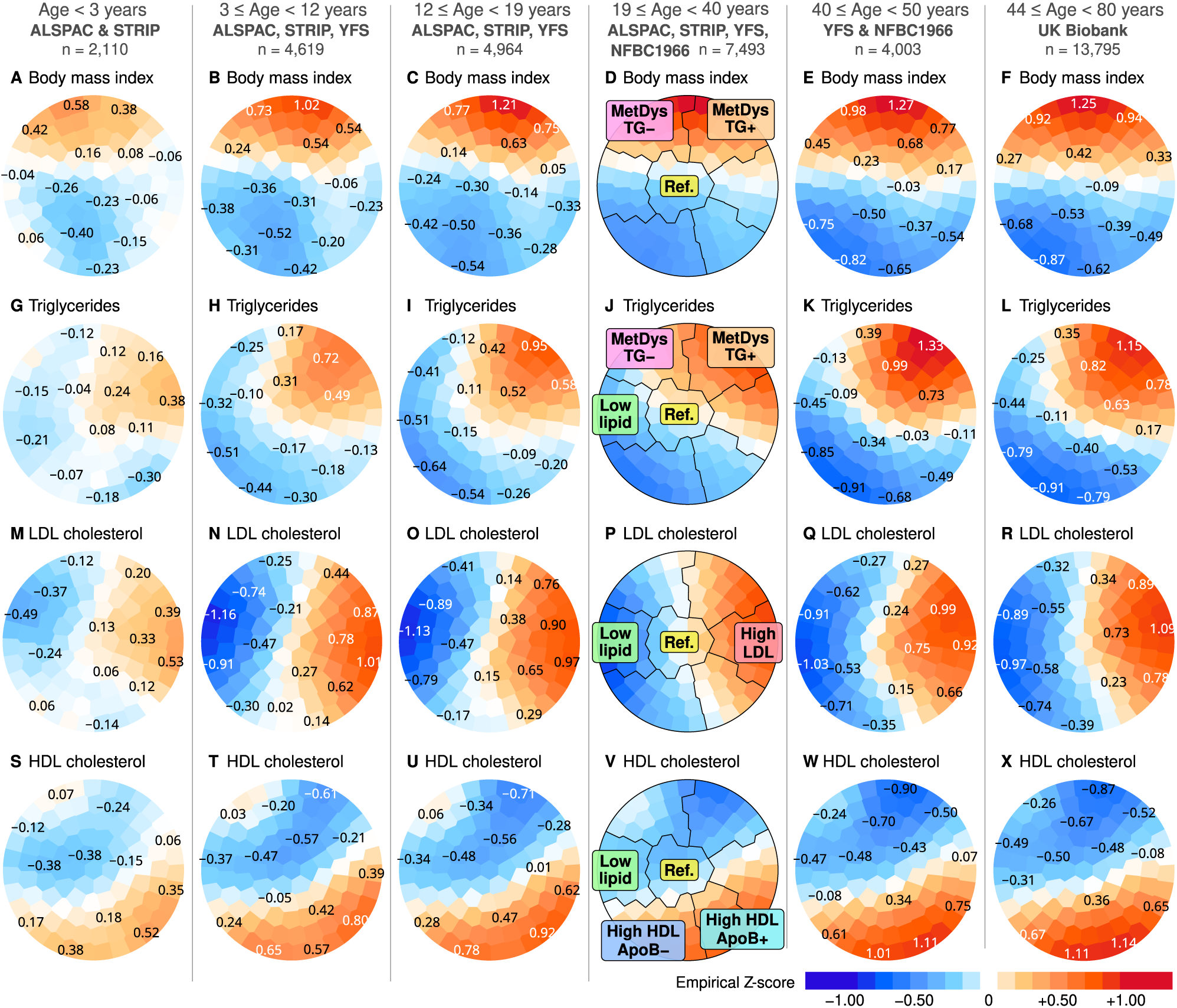
Temporal self-organizing map and the classification boundaries for metabolic subgroups. The similarity of the patterns between age strata indicates stable population stratification over the human life span. The model was trained with longitudinal data that included 146 metabolic variables from four cohorts that covered age segments up to 49 years. UK Biobank longitudinal data were projected onto the pre-trained model and the map colourings for measurements up to the age of 79 years are included in the figure (Plots F,L,R,X). Each coloured plot depicts the same map, that is, a two-dimensional layout of participants. Each participant is located in the same position of the map in each plot, while the plot is colored according to the regional averages of the residents when observed at a specific age segment (i.e. blue/red indicates lower/higher than population average). Participants were subsequently classified into seven subgroups by dividing the map into regions based on visual inspection of the colour patterns (Plots D,J,P,V). Abbreviations: Avon Longitudinal Study of Parents and Children (ALSPAC), apolipoprotein B (ApoB), high-density lipoprotein (HDL), low-density lipoprotein (LDL), metabolic dysfunction (MetDys), Northern Finland Birth Cohort 1966 (NFBC1966), Special Turku Coronary Risk Factor Intervention Project (STRIP), triglycerides (TG) and Cardiovascular Risk in Young Finns Study (YFS).

Individuals with high body mass index (i.e. potential metabolic dysfunction) were mostly located on the top half of the map and they were designated into two subgroups depending on the amount of triglycerides (‘MetDys TG−’ and ‘MetDys TG+’, Figure 2D,J). The MetDys subgroups also exhibited high insulin, glucose, branched-chain amino acids and the liver enzyme alanine amino transferase (Supplementary Figure S7). Going clockwise, the right-most sector of the map was characterized by high LDL cholesterol (‘High LDL’, Figure 2P) and the highest apolipoprotein B in older individuals (Supplementary Figure S7). HDL cholesterol was higher on the bottom half of the map, and we split the region further into two subgroups based on the concentration of apolipoprotein B (‘High HDL ApoB−’ and ‘High HDL ApoB+’, Figure 2P,V; see also apolipoproteins B and A-I in Supplementary Figure S7). The left-most sector of the map was characterized by low concentrations of all clinical lipids (‘Low lipid’, Figure 2J,P,V). Lipoprotein (a) was close to the population mean across all subgroups (Supplementary Figure S7).

### Metabolic ageing trajectories

Body weight and waist-height ratio were stratified early in life (Figure 3A,B). Weight stabilized by the age of 40 years, but waist continued to widen (slowly) into older ages (Supplementary Figure S8). Circulating insulin increased substantially in the two subgroups with high weight from the fourth decade onward, while there was a decrease from the puberty peak to pre-pubertal range in the High HDL ApoB− subgroup (Figure 3D). Weight stratification was correlated with C-reactive protein (Figure 3E), particularly in young women with high weight who showed higher concentrations than their male peers (Supplementary Figure S8).

**Figure 3:**
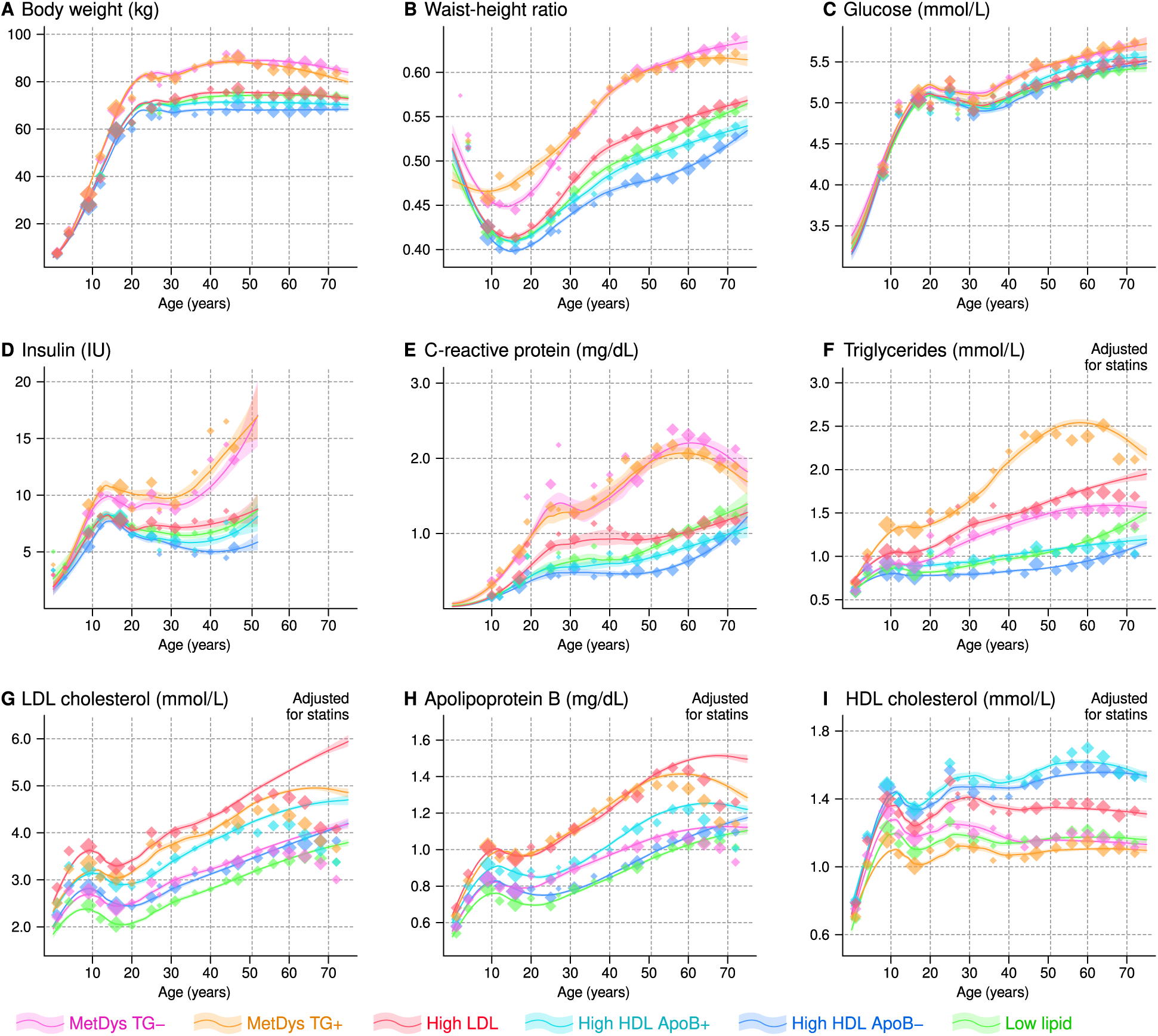
Trajectories of selected metabolic measures adjusted for cohort and batch effects. The markers indicate median adjusted values for age segments spaced at regular intervals. The biomarker curves and 95% confidence intervals were determined by local polynomial regression based on data that were further adjusted for statins and anti-hypertensive medication. Abbreviations: apolipoprotein B (ApoB), high-density lipoprotein (HDL), low-density lipoprotein (LDL), metabolic dysfunction (MetDys) and triglycerides (TG).

Triglyceride concentrations were the highest, by definition, in the MetDys TG+ subgroup (Figure 3F), and the trajectory stratification for other lipoprotein biomarkers was also consistent with the subgroup definitions (Figure 3G-I). Statin medication was associated with age and had a substantial impact on the observed LDL cholesterol trajectories (Figure 3G). The adjusted models suggest that LDL cholesterol would increase at a nearly consistent rate throughout most of adulthood in most participants in the absence of interventions.

The trajectory analyses indicated that the relative stratification within a peer group remained stable over decades. For this reason, we summarized the overall lipoprotein subclass and other biomarker stratification by lifetime z-scores (Supplementary Data).

### Cardiometabolic disease occurrence

We focused on the most common clinical classifications for obesity, diabetes and cardiovascular disease in the UK Biobank. At baseline, the majority of participants were free of recorded diagnoses (Figure 4 and Supplementary Table S4). The highest rates of (historical) diagnoses were observed for the two high-weight subgroups (Figure 4A-C). Notably, men in the MetDys TG− had the highest diabetes prevalence (10.2%; CI95: 8.6%, 12.0%).

**Figure 4:**
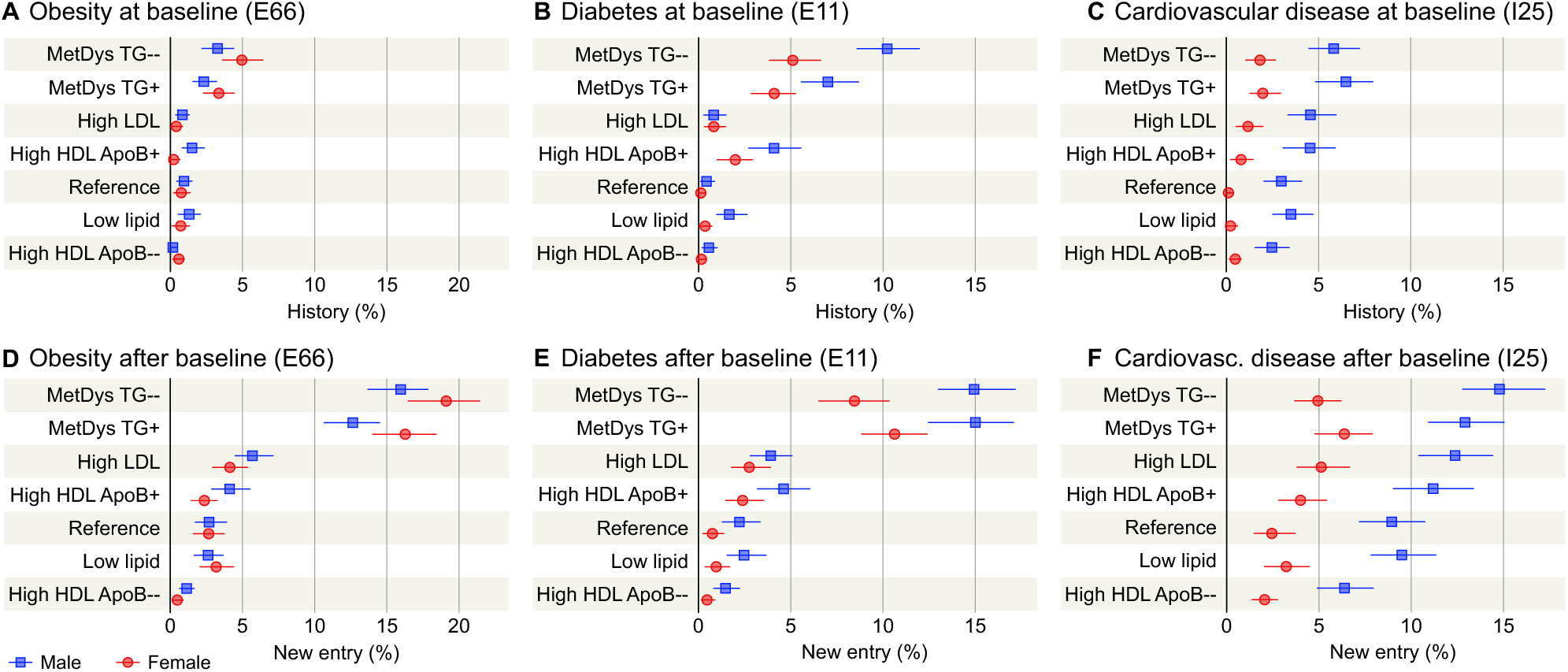
Prevalence and incidence of common cardiometabolic diagnoses in the UK Biobank (n = 13,795) by metabolic subgroup. The mean follow-up time was 14.5 years (Plots D-F). Diagnosis codes are from the International Statistical Classification of Diseases and Related Health Problems, 10th Revision. Abbreviations: apolipoprotein B (apoB), high-density lipoprotein (HDL), low-density lipoprotein cholesterol (LDL) and triglycerides (TG).

The patterns for new entries during a mean follow-up of 14.5 years were similar to the retrospective data at baseline but more pronounced (Supplementary Table S5). Obesity was recorded in over 15% of women in the two MetDys subgroups but for less that 5% of women in other subgroups (Figure 4D). Incident diabetes in men occurred in 14.9% (CI95: 13.0%, 17.2%) of the MetDys TG− subgroup members, while the incidence was less than 5% outside the MetDys groups (Figure 4E). We observed substantial divergence between men and women with respect to cardiovascular disease: the incidence was 14.8% (CI95: 12.8%, 17.2%) for men in the MetDys TG− subgroup, but only 4.9% (CI95: 3.7%, 6.2%) for their female peers with the same metabolic classification (Figure 4F).

Lastly, we compared the relative risk of cardiometabolic diagnoses against the lowest risk category (High HDL ApoB−) after adjusting for baseline age and sex (Table 1). The highest hazard ratios for diabetes were observed for the MetDys TG− (HR = 13.24; CI95 8.54, 20.52) and MetDys TG+ (HR = 15.03; CI95 9.73, 23.23) subroups. Similarly, the highest hazard ratios for cardiovascular disease were observed for the MetDys TG− (HR = 2.54; CI95 1.99, 3.25) and MetDys TG+ (HR = 2.56; CI95 2.00, 3.28) subroups.

**Table 1:** Hazard ratios of incident clinical classifications for metabolic subgroups. A Cox regression model was fitted for each clinical code in the longitudinal subset of the UK Biobank (n = 13,795, 14.5 years of follow-up). Metabolic subgroup labels were encoded into six binary regressors; the High HDL ApoB− was assigned as the “no risk” subgroup and thus not included as a regressor due to redundancy. The models were adjusted for sex and baseline age. The numbers in brackets indicate the 95% confidence interval. Diagnosis codes are from the International Statistical Classification of Diseases and Related Health Problems, 10th Revision.

|  | MetDys TG– | MetDys TG+ | High LDL | High HDL ApoB+ | Reference | Low lipid | High HDL ApoB– |
| --- | --- | --- | --- | --- | --- | --- | --- |
| **Diabetes (E11)** | 13.24<br>(8.54, 20.52) | 15.03<br>(9.73, 23.23) | 3.25<br>(2.01, 5.26) | 3.61<br>(2.21, 5.90) | 1.73<br>(0.99, 3.05) | 1.79<br>(1.03, 3.10) | 1.00 |
| **Obesity (E66)** | 25.21<br>(15.69, 40.50) | 20.58<br>(12.79, 33.11) | 6.41<br>(3.88, 10.59) | 4.16<br>(2.44, 7.09) | 3.69<br>(2.14, 6.36) | 3.80<br>(2.22, 6.52) | 1.00 |
| **Lipoprotein disorder (E78)** | 1.65<br>(1.38, 1.97) | 2.98<br>(2.52, 3.51) | 3.16<br>(2.68, 3.74) | 1.81<br>(1.51, 2.18) | 1.70<br>(1.41, 2.03) | 0.85<br>(0.69, 1.05) | 1.00 |
| **Hypertension (I10)** | 2.47<br>(2.10, 2.91) | 2.34<br>(1.99, 2.75) | 1.61<br>(1.37, 1.90) | 1.45<br>(1.21, 1.73) | 1.12<br>(0.93, 1.35) | 1.27<br>(1.06, 1.51) | 1.00 |
| **Ischemic heart disease (I25)** | 2.54<br>(1.99, 3.25) | 2.56<br>(2.00, 3.28) | 2.01<br>(1.57, 2.59) | 1.81<br>(1.39, 2.37) | 1.59<br>(1.20, 2.11) | 1.54<br>(1.17, 2.04) | 1.00 |

## Discussion

To our knowledge, this is the first epidemiological study that integrates comprehensive biochemical time-series from birth to old age. Multivariable subgroup trajectory modelling elucidated the temporal patterns of cardiometabolic health across seven decades of life and we connected the subgroups with cardiometabolic morbidity up to the ninth decade of life. We revealed a stable cardiometabolic stratification from childhood to adulthood that correlates with divergent disease incidence. Accumulation of body weight from the second to the fifth decade of life was a key feature of the stratification and may represent a critical period for public health interventions. From a statistical perspective, central obesity combined with molecular ageing and lipoprotein metabolism explained the population diversity in cardiometabolic trajectories and associated clinical events.

Young children have low cholesterol, then an increase before puberty, followed by a modest decline towards the end of the second decade ([29] and references therein). In adults, cholesterol increases until the fifth decade after which a plateau is observed first in men and then a decade later in women [32–34]. LDL cholesterol shows the same general trend as total cholesterol, and both typically decrease in the oldest age groups. Our trajectory modelling replicated the aforementioned broad patterns, except for the statin-adjusted model for LDL-cholesterol that did not show the expected decline in all older participants (Figure 3G). The old age decline was reported before statins became available [35], and therefore the discrepancy may be due to the lack of compatibility regarding the literature-based adjustments [28]. Without the adjustments, the statin effect reduced subgroup differences in atherogenic lipids (note how the spread between observations decreases in old age in Figure 3G,H). This may partly explain why the two high weight subgroups in experienced comparable or even higher cardiovascular disease rates than the High LDL subgroup that would be expected to show the highest risk [36]. We suspect that after equalizing the causal lipoproteins, obesity- and diabetes-related competing causes may have become more important [37].

Glucose metabolism deteriorates with age [38] and the consistent increase in circulating glucose we saw in adulthood fits with previous reports [39,40]. Similarly, chronic inflammation is associated with ageing hallmarks [41] and the detailed temporal trends from this study are compatible with earlier reports [42,43]. Of note, the higher C-reactive protein in young women (Supplementary Figure S8) can be attributed to the menstrual cycle and hormonal contraception [44–46]. Both glucose and insulin peak during puberty [29], and this study revealed that insulin diverged in adulthood depending on the metabolic subgroup while stratification in circulating glucose remained modest, even late in life (Figure 3C,D). Although the insulin trajectories end before age 50 (not available in the UK Biobank), they nevertheless predicted diabetes in the UK Biobank via the subgroup profiles (Figure 4E). This reflects the likely etiological process where obesity-associated insulin resistance leads to higher insulin demand and beta-cells stress in genetically susceptible individuals [38,47]. Eventually, the growing gap between demand and capacity results in hyperglycemia that exceeds clinical diabetes criteria.

The causal and observational associations between obesity and human health have been extensively documented [48–50] and there is a growing interest in molecular subclassification of obesity and how it predicts specific diseases [11,12,51]. The data patterns we observed led us to define two subgroups with high average weight (MetDys) that were segregated by circulating triglycerides (Figure 2D). Surprisingly, the subdivision was not associated with a substantial difference in morbidity as one would expect by the tight association between triglycerides and hyperglycemia. More frequent monitoring and aggressive statin, blood pressure and other preventative treatments in the TG-rich high weight subgroup would be typical under the metabolic syndrome umbrella [52]. Therefore, the disease risk for MetDys TG− individuals may be under-appreciated by current guidelines due to their more favourable lipoprotein profile.

Obesity without metabolic dysfunction has been proposed as an important clinical entity [12]. These individuals were present in our study – there were obese phenotypes in every subgroup, albeit at varying proportions (Figure 4A,D). From a technical perspective, the residual body mass distribution around the subgroup mean is always going to have an upper and lower tail, which explains the non-zero prevalence. At subgroup level, “healthy obesity” will not manifest unless the combination of high weight and low-risk biochemistry is frequent; we did not see evidence for such a combination, which means that these individuals were rare. In cross-sectional studies, incidental combinations of obesity and low-risk biochemistry may produce a statistical signal but such signals were reduced by the temporal scope of our study. Previous longitudinal studies suggest that healthy obesity does not stay healthy in the long term [53,54], which may explain the rarity of these individuals in our life course study.

The inclusion of multi-cohort longitudinal data and large sample sizes are strengths of this study, and we also conducted careful statistical verification that the observed patterns were not methodological artefacts. On the other hand, temporal data coverage was incomplete, thus further studies with independent longitudinal data are warranted to confirm the findings, especially considering the narrow ethnic focus on North European populations. Furthermore, metabolomics data coverage was insufficient for reliable trajectory modelling, which limited the molecular scope of the study. The input data included more lipoprotein lipids than other types of molecules (hence the lipoprotein-based subgroups) but, as lipoprotein lipids are clinically important, we do not think this would invalidate the take-home messages. This is an observational study, thus we caution against causal or too mechanistic interpretation of our findings unless already established in the literature.

Obesity poses a general risk to health [55], and the importance of early prevention of metabolic dysfunction cannot be underestimated. We identified the age range between 9 and 40 years as the critical period when most of the divergence in body mass occurred and the ages between 17 and 30 when the paths towards hyperinsulinemia and diabetes were still in tandem before an exponential increase in the MetDys subgroups during the fifth decade. Importantly, the population structure that underlies these patterns was detectable in pre-pubertal children (Figure 2B). The STRIP study pioneered lifestyle interventions focused on cardiometabolic risk factors with a detectable effect on childhood obesity [56]. Our trajectory analyses indicate that weight gain early in life may represent a major target for prevention in contemporary populations since the impact is likely to be substantially broader than just LDL dyslipidaemia or other specific causes. GLP-1 receptor antagonists have already been trialled in children [16]. Therefore, we are optimistic that new public health programs that combine traditional advice for healthier lifestyle with pharmacological support may be able to reverse the global trend of increasing obesity-associated metabolic dysfunction.

## Ethics approval

This study did not involve recruitment of study subjects or new biomedical experiments. Ethical approval for ALSPAC was obtained from the ALSPAC Ethics and Law Committee and the Local Research Ethics Committees. The STRIP was approved by the joint Committee of Ethics of the University of Turku and the University Central Hospital of Turku on Nov 8th 1989. The YFS was approved by the ethical committees for five Finnish universities with medical schools (Regional Committee on Medical Research Ethics of the HUS Joint Authority for Helsinki, Regional Medical Research Ethics Committee of Eastern Finland for Kuopio, Northern Ostrobothnia Hospital District Ethical Committee for Oulu, the Ethics Committee of the Wellbeing Services County of Pirkanmaa for Tampere and the Ethics Committee of the Wellbeing Services County of Southwest Finland for Turku). The NFBC1966 study was approved by the Northern Ostrobothnia Hospital District Ethical Committee, Finland (94/2011, Dec 12^th^ 2011). The UK Biobank was approved by the North West Multi-centre Research Ethics Committee (21/NW/0157, Jun 29^th^ 2021) with details available online [https://www.ukbiobank.ac.uk/learn-more-about-uk-biobank/about-us/ethics].

For ALSPAC, consent for biological samples has been collected in accordance with the Human Tissue Act (2004). Informed consent for the use of all data collected was obtained from participants following the recommendations of the ALSPAC Ethics and Law Committee at the time. Participants can contact the study team at any time to retrospectively withdraw consent for their data to be used. Study participation is voluntary and during all data collection sweeps, information was provided on the intended use of data. Specific Research Ethics Committee approval is sought for the consenting process at each collection sweep. Written consent, including permission for future use, is obtained from adult participants or from the parents of children as appropriate. Ethical approval for future use is covered by ALSPAC’s Research Tissue Bank approval. All historical consents to hold biological samples have been reviewed as part of the Tissue Bank approval process. Participants can contact the study team at any time to retrospectively withdraw consent for use of their samples. For STRIP, written informed consent was obtained from parents at study entry, and from the participants at ages 15, 18, and 26 years. For YFS, NFBC1966 and UK Biobank, written informed consent was also obtained.

## Data availability

The ALSPAC data is available to researchers via application (URL: https://www.bristol.ac.uk/alspac/). This study was approved under the project B3830. The Finnish datasets used in the current study are available from the cohorts through application process for researchers who meet the criteria for access to confidential data (URL: http://youngfinnsstudy.utu.fi and URL: https://stripstudy.utu.fi/). Regarding these data, the ethics committee has concluded that under applicable law, the data from these studies cannot be stored in public repositories or otherwise made publicly available. The data controller may permit access on case-by-case basis for scientific research, not, however, to individual participant level data, but aggregated statistical data, which cannot be traced back to data for the individual participants. The UK Biobank data are available to the public (URL: https://www.ukbiobank.ac.uk/). This study was part of Project #324302.

## Author contributions

V-PM conceived and designed the study, did the statistical analyses, and interpreted the results. V-PM wrote the manuscript with help from MA-K. MK, TL, NH, TR, JV, KP, SR, HN, JM, OR and MA-K collected and/or produced data. All authors discussed the results, commented, and contributed to the manuscript, and approved the final version

## Use of artificial intelligence (AI) tools

The Elicit online service (URL:https://elicit.com/) was used for searching relevant literature. No other tools were used.

## Acknowledgements

We are extremely grateful to all the families who took part in this study, the midwives for their help in recruiting them, and the whole ALSPAC team, which includes data collection staff, data and administrations staff, technical managers and the technical staff with the Bristol Bioresource Laboratory, based within the University of Bristol. The STRIP study children and their parents and grandparents have made this study possible – the authors thank them for their time, continued efforts, and commitment to the STRIP throughout the years. We thank and acknowledge the participation and contributions from the individuals and their families of the YFS cohort through multiple decades of follow-up. We also thank and acknowledge the participation and repeated follow-up visits from the participants of the NFBC1966 and UK Biobank.

## Supplementary material

### Technical quality control of the subgroup analysis

In the first part of study, we investigated the longitudinal multivariable structure of cardiometabolic health using the self-organising map (SOM) algorithm. The first practical steps were to identify a suitable high-coverage subset for trajectory analyses (Figure S1) and to describe the basic characteristics of the cohorts for context (Figure S4). A substantial set of mostly cross-sectional data were left over, and these data were used to fix initial conditions and parameters for the subgroup modelling independently of the trajectory training set (Figure S1B).

We used the balancing feature implemented in the Numero R library (reference in the main text) to reduce the statistical effects from categorical confounders. The balance parameter of the SOM algorithm was set to 100%, which forced equal numbers of resident samples per map district in the final layout. Importantly, the balancing was applied separately for cohort and sex categories, thus eliminating their confounding effects.

For quality control, we confirmed that the final SOM (Figure 2 in main text) was not confounded by cohort or sex differences (Figure S5). We also created a randomized version of the training set where the cross-sectional associations between variables were preserved within age strata, but the longitudinal dependencies were removed. An alternative SOM was then trained with the selectively randomized inputs, while the initial map state and all other settings were copied from the original model. Side-by-side comparison of the results confirmed that the unaltered dataset produced a longitudinally consistent SOM while the randomized inputs did not (Figure 2 versus Figure S6). This indicated that the consistency between the temporal strata in Figure 2 was driven primarily by the temporal continuity within the serial measurements of a single individual.

---

**Figure S1:**
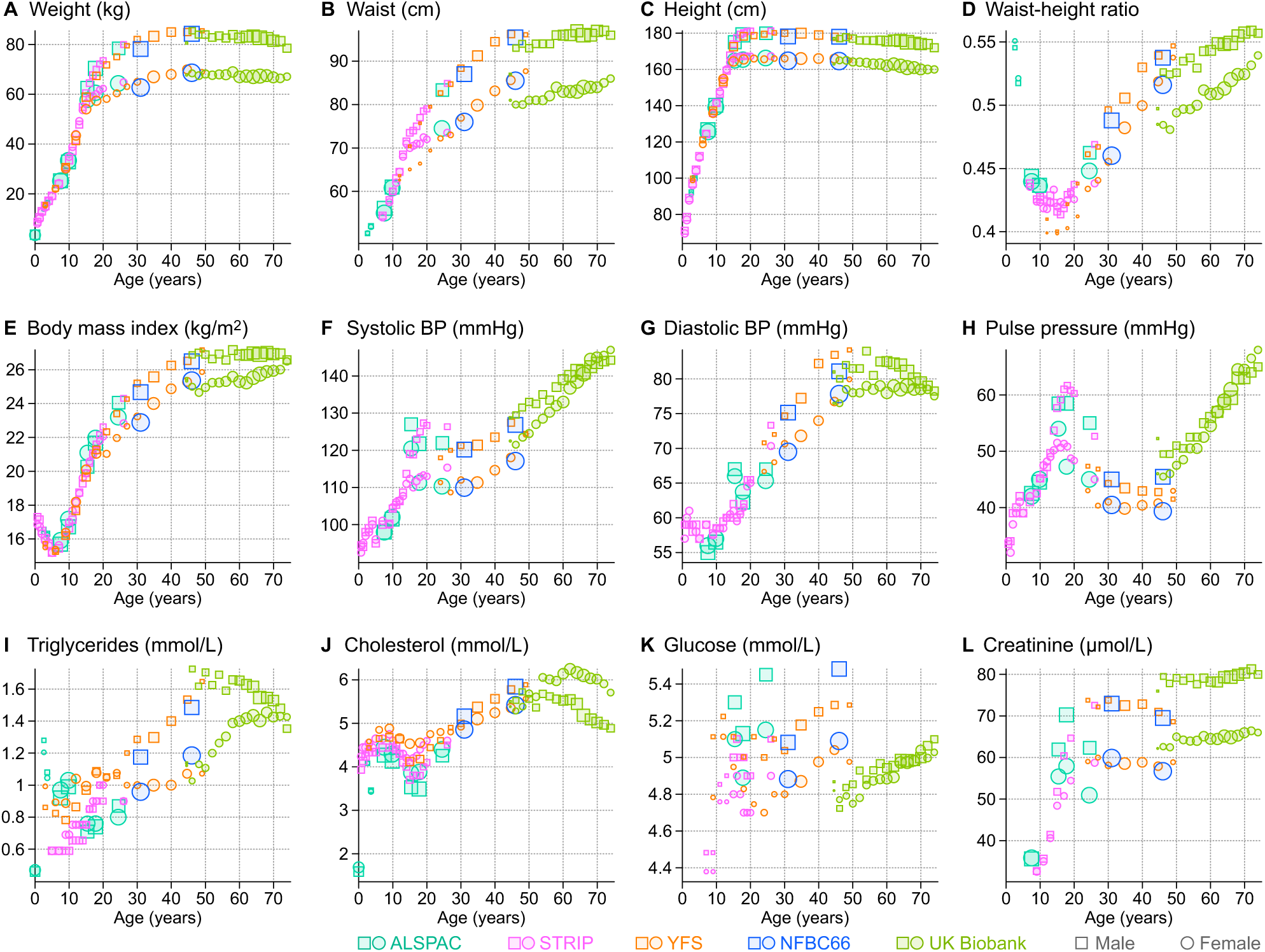
Overview of the study cohorts. Only participants with longitudinal data are included from the UK Biobank. The markers indicate median values for a given age stratum and their size indicates the number of data points. The strata were defined as rounded age (ALSPAC, STRIP, YFS and NFBC1966) or as two-year segments (UK Biobank).

**Figure S2:**
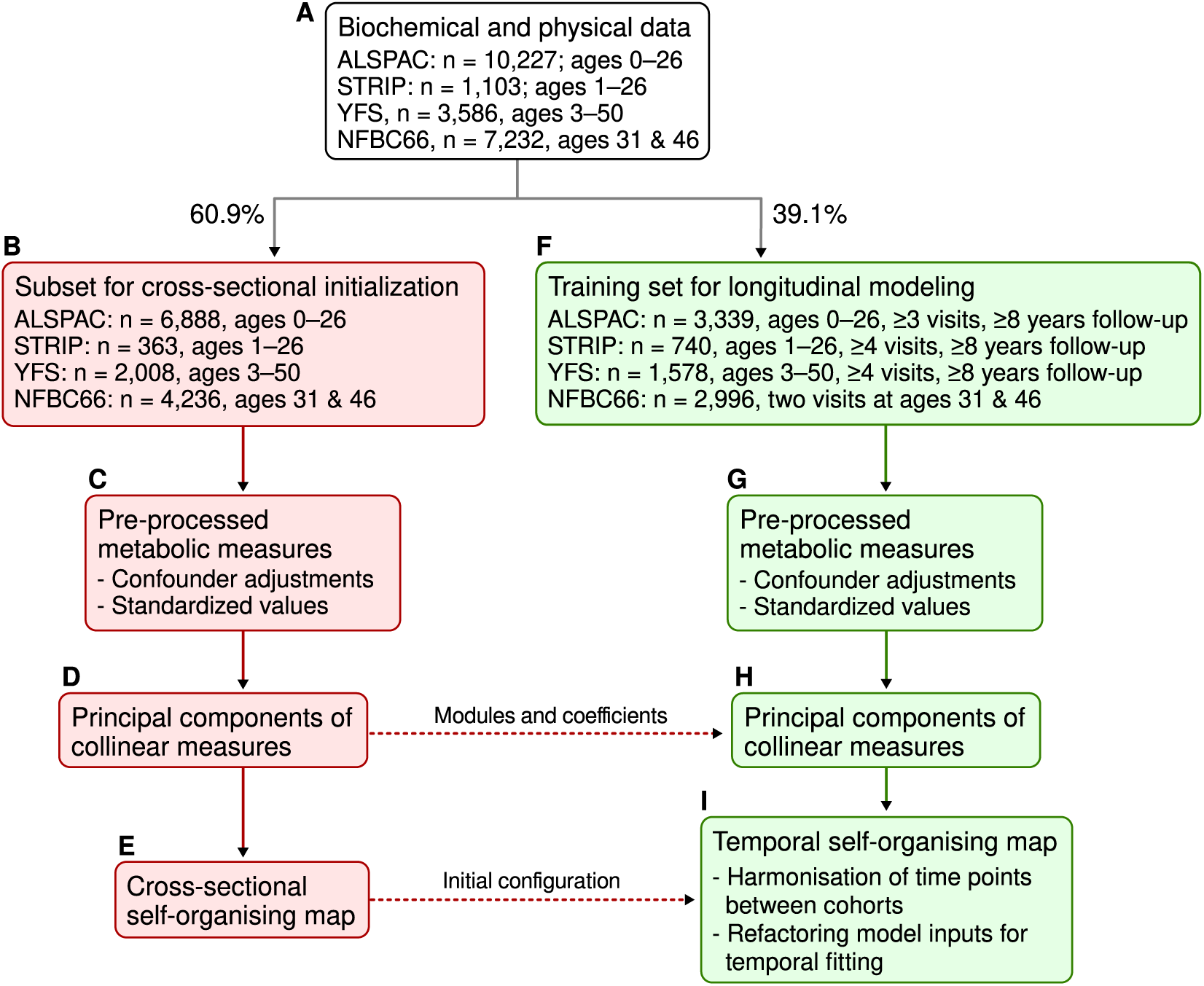
Selection of data for training the temporal self-organizing map.

**Figure S3:**
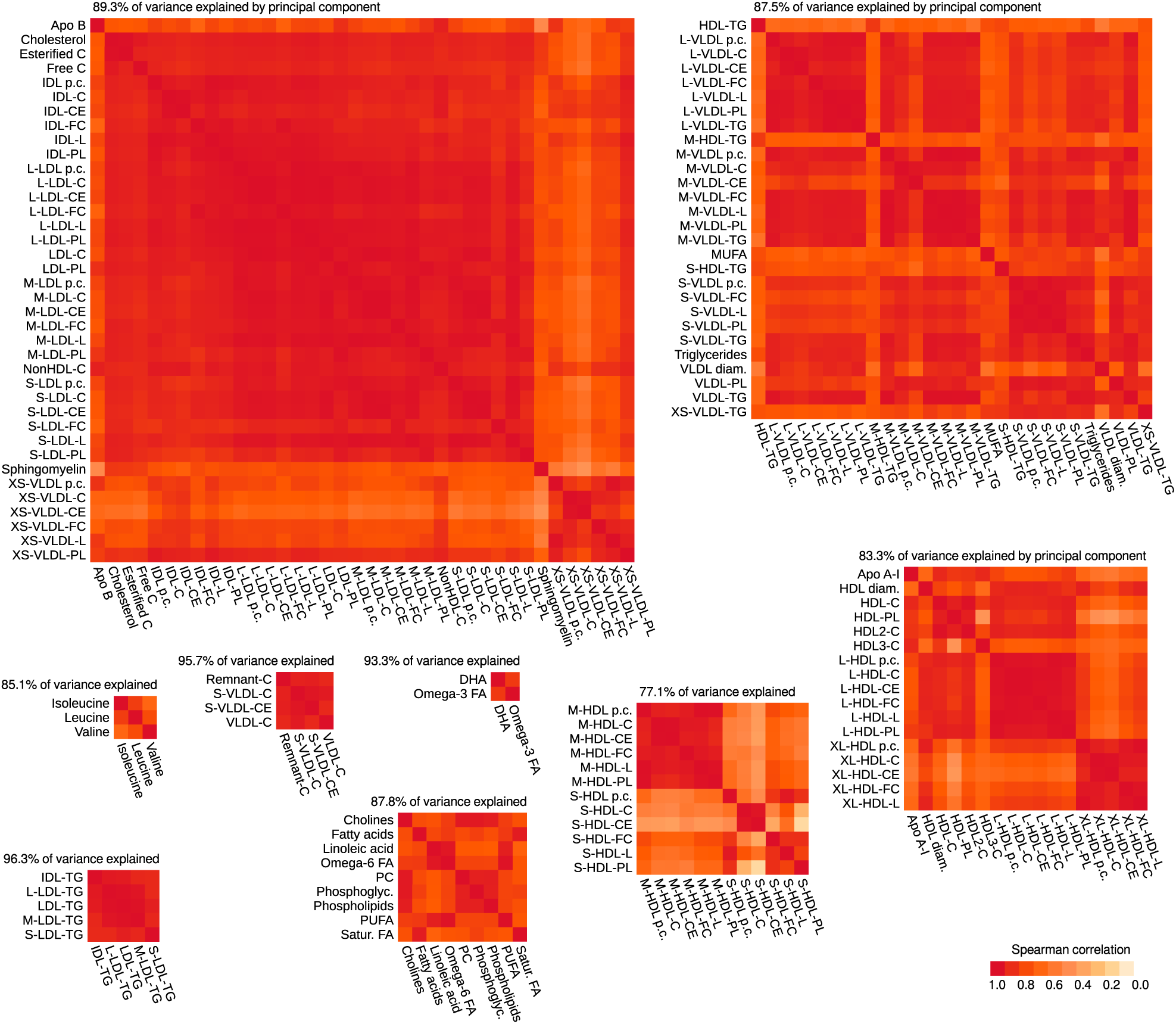
Merging of collinear NMR variables based on the cross-sectional subset. The principal component was calculated for each module as the summary score to be used in subsequent statistical analyses. The same component weights were applied to the longitudinal training set before fitting the temporal self-organizing map. Weigh coefficients for the variables are included in Supplementary Data.

**Figure S4:**
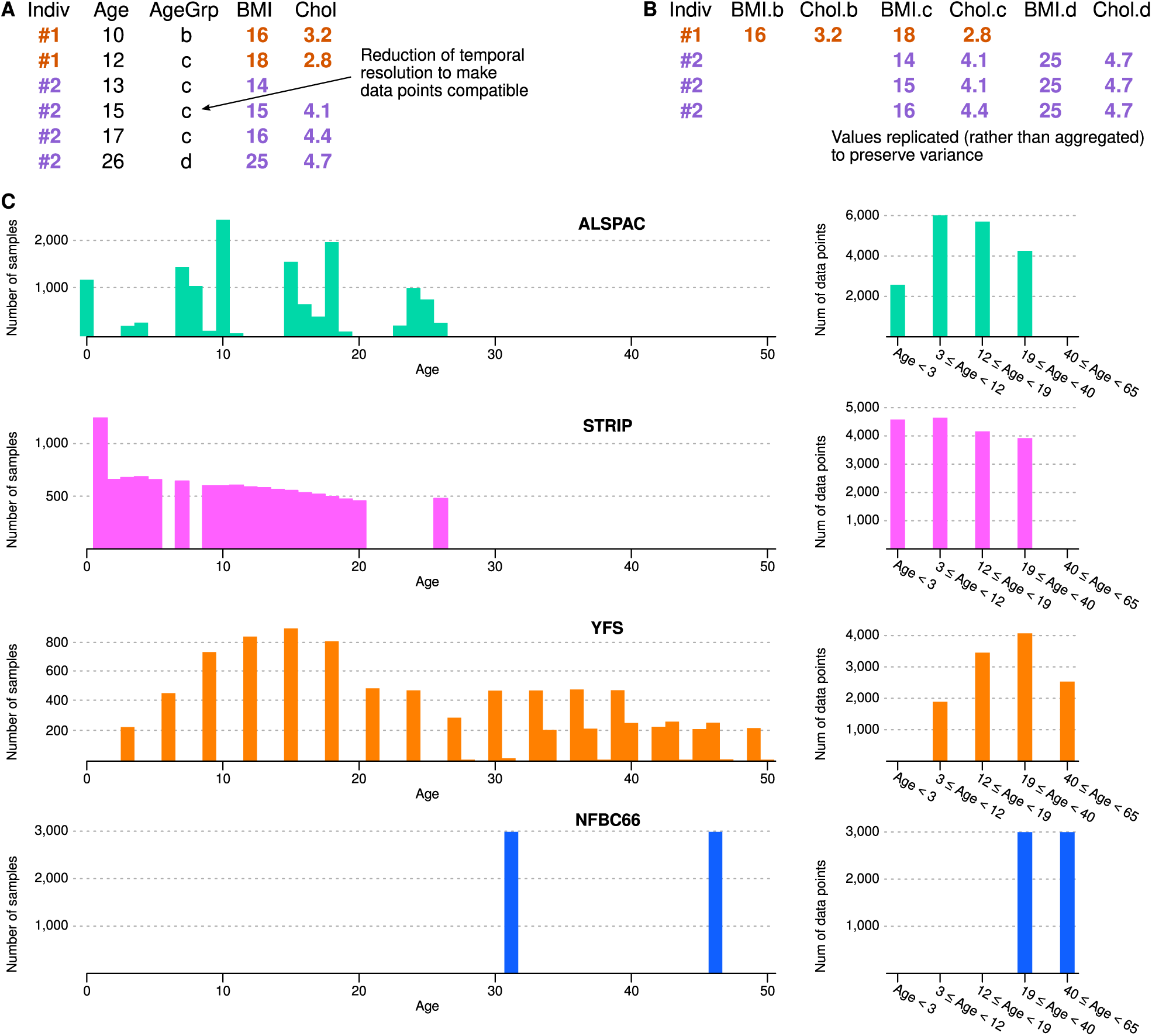
Harmonization of time points and refactoring of the data matrix for temporal self-organizing map. Plot A shows a schematic example of a longitudinal dataset with multiple age points per participant. In this study, the age points came from multiple cohorts with varying designs, thus we harmonized the ages by assigning each point to a wider peer group. We also tested an alternative strategy where parametric curves were fitted to each cohort separately and then resampled at compatible time points, however, this method failed to produce stable results due to substantial cohort differences (data not shown). Plot B shows the refactoring procedure where each variable (BMI and cholesterol in this example) was expanded to multiple columns such that a complete trajectory could be contained within a single row in the new matrix. For many participants, several time points were contained within the peer group, however, it was not possible to aggregate these as that would have altered the individual variance of the data, and would have led to artificial clustering on the SOM. Instead, we used an imputation scheme to reconstruct complete trajectories by replicating values as needed. Consequently, individuals with dense time series were represented by multiple rows in the final training set. The duplication of data values explains the difference between the number of samples (Plot C, left side) and the final data points for training (Plot C, right side). Of note, STRIP includes two visits that both round up to one year, which explains the high number of early life samples.

**Figure S5:**
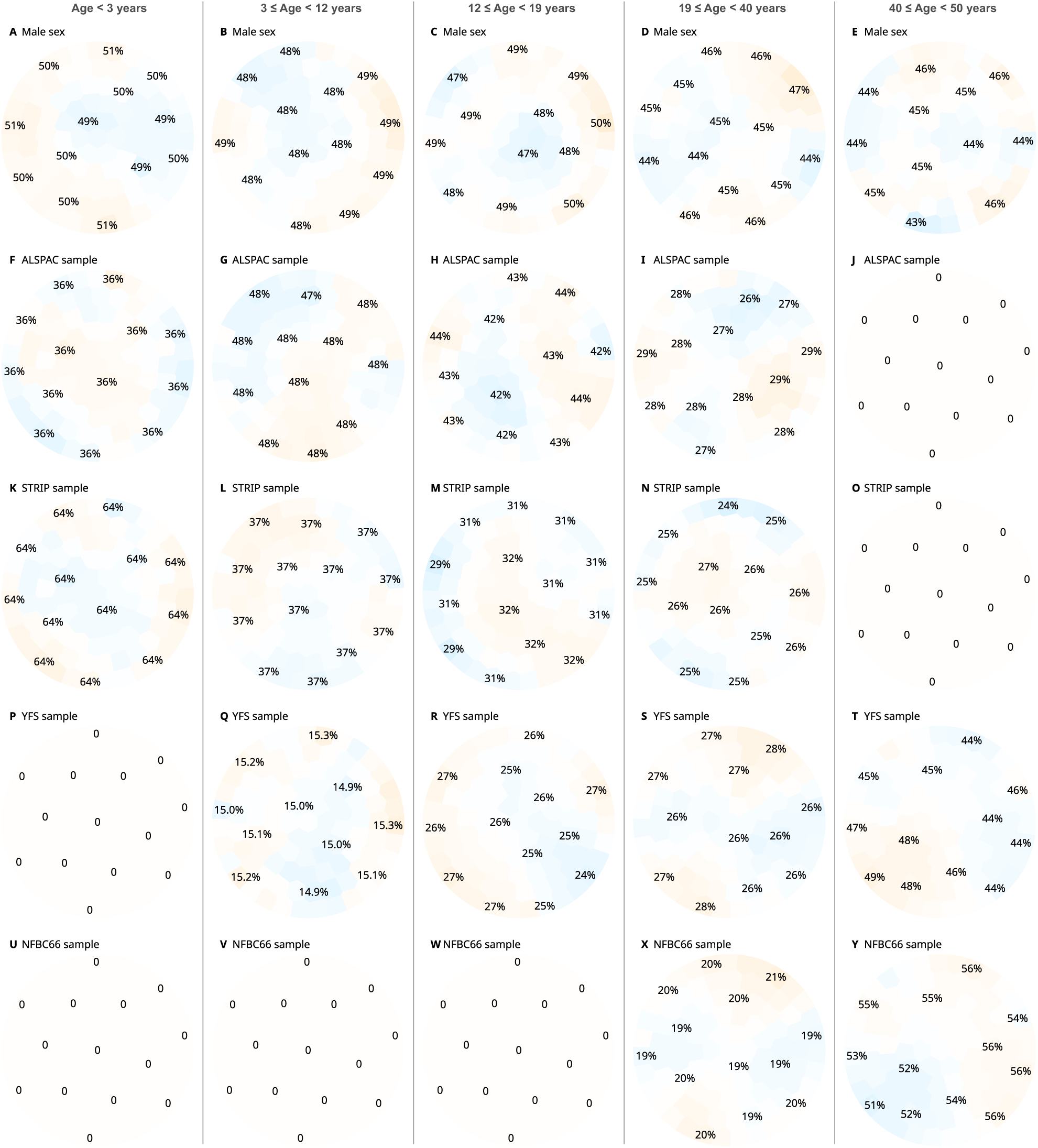
SOM colorings for confounding categories. The numbers on the plots indicate the proportion of a specific category in a region. The total number of data points that includes all cohorts within an age group is shown under the plot titles. The map was trained with the balance parameter set at 100%. This means that during each training cycle, the locations of data points are optimized such that each map district ends up with the same number of residents. For the final layout, locations were determined in batches so that the distribution of individuals of the same sex and same cohort was as uniform as possible across the map and age groups.

**Figure S6:**
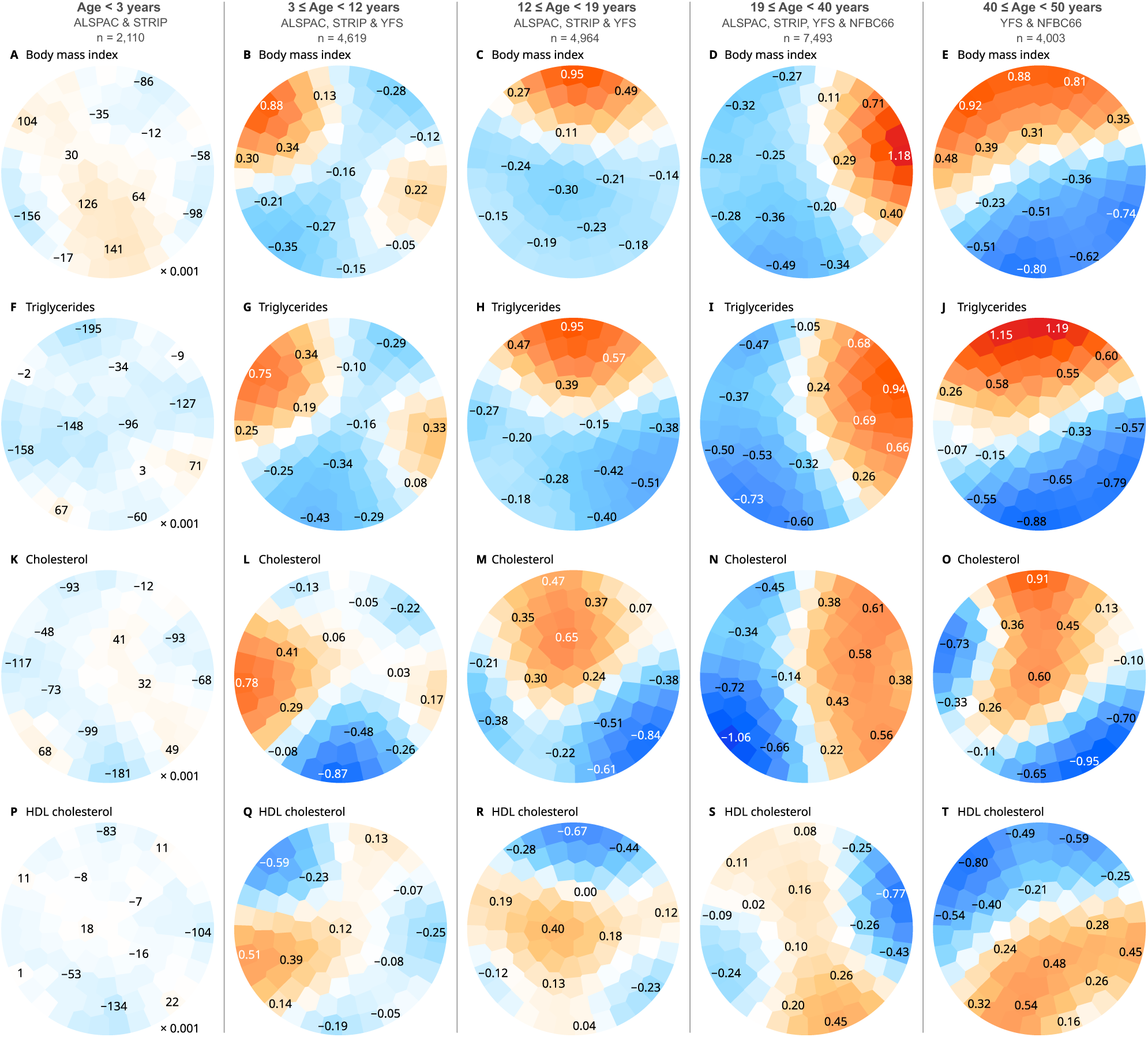
An alternative SOM trained with randomized longitudinal information. Unlike the SOM trained with the unaltered data (Figure 2 in main text), the lack of longitudinal links between data points led to fragmented regional patterns where the cross-sectional relationships were preserved within an age group, but the patterns were not consistent between the age groups. For example, body mass index and triglycerides were increased in the same regions in young children aged between 3 and 12 years (Plots B and G). In parallel, body mass index and triglycerides show the same cross-sectional relationship in adults between 19 and 40 (Plots D and I), but there is no regional continuity between the age groups.

**Figure S7:**
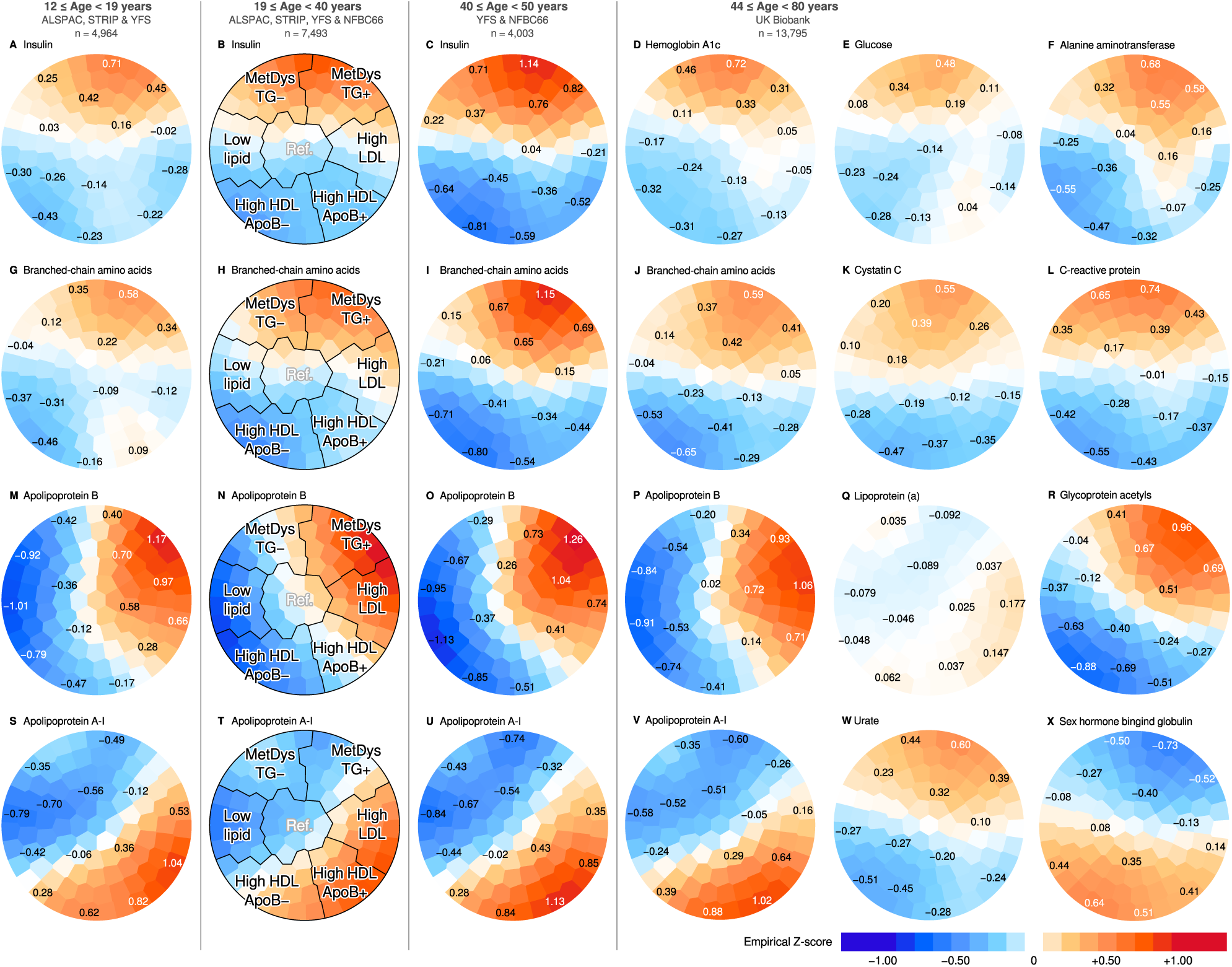
Additional SOM colorings for biochemical variables, including clinical biomarkers in the UK Biobank that were not available from the other cohorts. Empirical z-scores were calculated by splitting the original data by age, sex and cohort membership into batches, and then the data within each batch was standardized to zero mean and unit variance.

**Figure S8:**
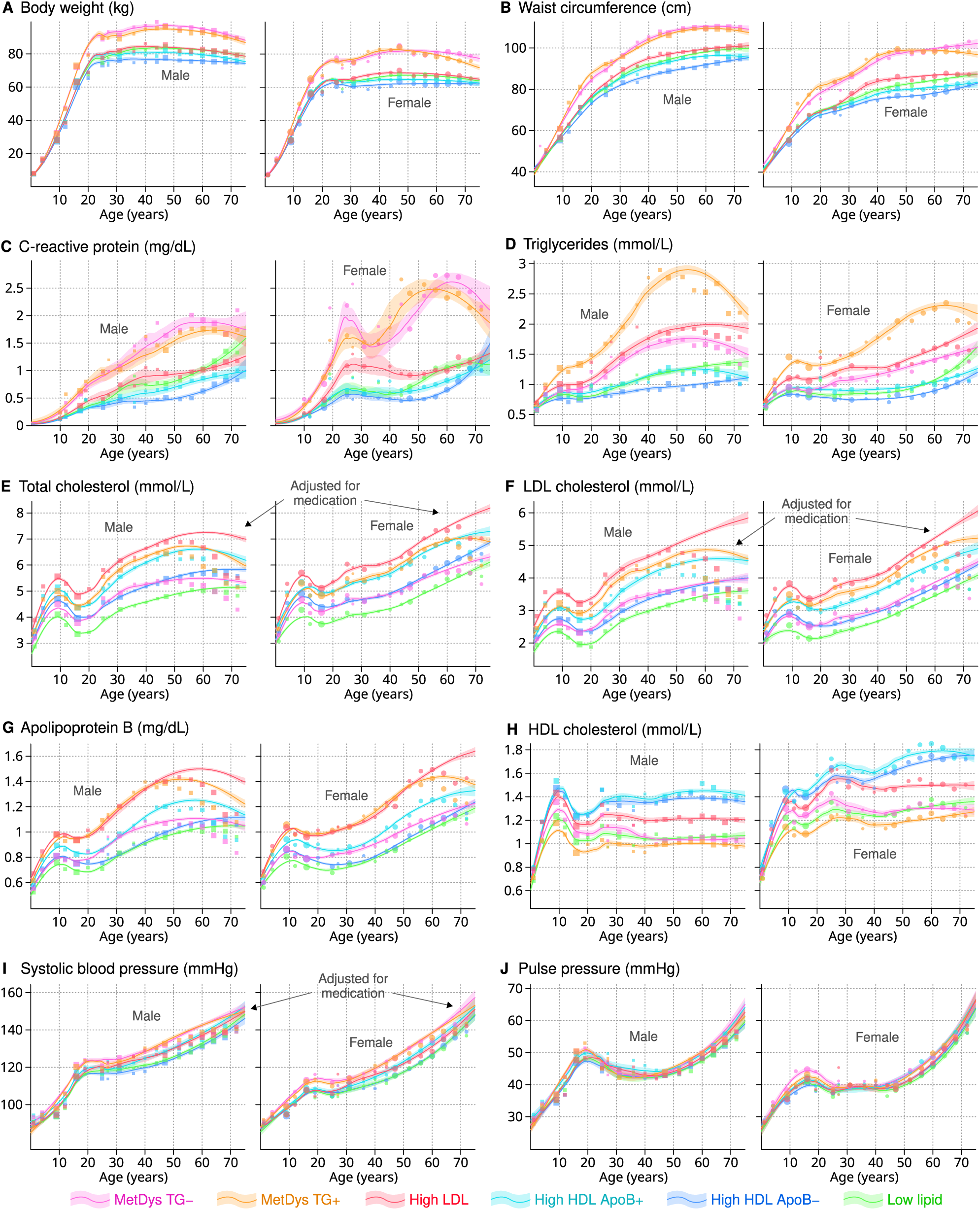
Longitudinal trajectories of metabolic measures adjusted for cohort differences and medications. The curves were modelled by polynomial local regression (adjusted for medication). Squares and circles indicate the medians for male and female participants, respectively (not adjusted for medication). The size of the markers reflects the number of data points that were used for calculating the aggregate estimate.

**Table S1:** Cohort descriptions.

|  |  |
| --- | --- |
| ALSPAC | The Avon Longitudinal Study of Parents and Children is a birth cohort from the South West of England. Pregnant women resident in Avon, UK with expected dates of delivery between 1st April 1991 and 31<sup>st</sup> December 1992 were invited to take part in the study. 20,248 pregnancies have been identified as being eligible and the initial number of pregnancies enrolled was 14,541. Of the initial pregnancies, there was a total of 14,676 fetuses, resulting in 14,062 live births and 13,988 children who were alive at 1 year of age. When the oldest children were approximately 7 years of age, an attempt was made to bolster the initial sample with eligible cases who had failed to join the study originally. As a result, when considering variables collected from the age of seven onwards (and potentially abstracted from obstetric notes) there are data available for more than the 14,541 pregnancies mentioned above: The number of new pregnancies not in the initial sample (known as Phase I enrolment) that are currently represented in the released data and reflecting enrolment status at the age of 24 is 906, resulting in an additional 913 children being enrolled (456, 262 and 195 recruited during Phases II, III and IV respectively). The phases of enrolment are described in more detail in the cohort profile paper and its update. The total sample size for analyses using any data collected after the age of seven is therefore 15,447 pregnancies, resulting in 15,658 fetuses. Of these 14,901 children were alive at 1 year of age. Of the original 14,541 initial pregnancies, 338 were from a woman who had already enrolled with a previous pregnancy, meaning 14,203 unique mothers were initially enrolled in the study. As a result of the additional phases of recruitment, a further 630 women who did not enrol originally have provided data since their child was 7 years of age. This provides a total of 14,833 unique women (G0 mothers) enrolled in ALSPAC as of September 2021. The study website contains details of all the data that are available through a fully searchable data dictionary and variable search tool (URL:http://www.bristol.ac.uk/alspac/researchers/our-data/). |
| STRIP | The Special Turku Coronary Risk Factor Intervention Project was a randomized controlled trial to investigate if a favourable diet would improve cardiometabolic risk factors. The intervention was initiated in 1990 and continued until age 20. Initial recruitment was 1,880 infants aged 5 months, of whom 1,062 went through randomization and 532 remained at age 14. In this study, biochemical data collected up to age 26 were included (standard biochemistry: n = 450–1,103, 20 time points; metabolomics: n = 444–803, 7 time points). |
| YFS | The Cardiovascular Risk in Young Finns Study recruited 3,596 children across six peer groups in 1980 in five cities in Finland. In this study, biochemical data measured up to 50 years of age were used. Clinical lipids were available from age 3 onward and both metabolomics and clinical assays were available from three visits in 2001 (1,239 women and 1007 men), 2007 (1,186 women and 974 men) and 2011 (1,112 women and 927 men). |
| NFBC1966 | The NFBC1966 is a longitudinal birth cohort established to study factors affecting preterm birth and consequent morbidity in the two northern- most provinces of Finland, Oulu and Lapland. The NFBC1966 includes 12,231 births (12,058 alive) covering 96% of all eligible births in this region during January–December 1966. Data collections in 1997 (at age of 31) and 2012 (age 46) including clinical examination and fasting serum sampling was used in the present study. Metabolomics and clinical assays were available from the 31-year (2,962 women and 2749 men) and 46-year visits (3,237 women and 2,549 men). |
| UK Biobank | The UK Biobank is a prospective cohort study of 502,148 participants aged 37–73 years recruited between 2006 and 2010. Polygenic risk scores for common traits were obtained from the biobank database (Categories 301 and 302). Disease outcomes were obtained through register linkage, including Hospital Episode Statistics (HES), cancer and national death registries (Category 1712, accessed in the first half of 2025). 13,795 participants with follow-up biochemistry data were included in this study. |

**Table S2:**
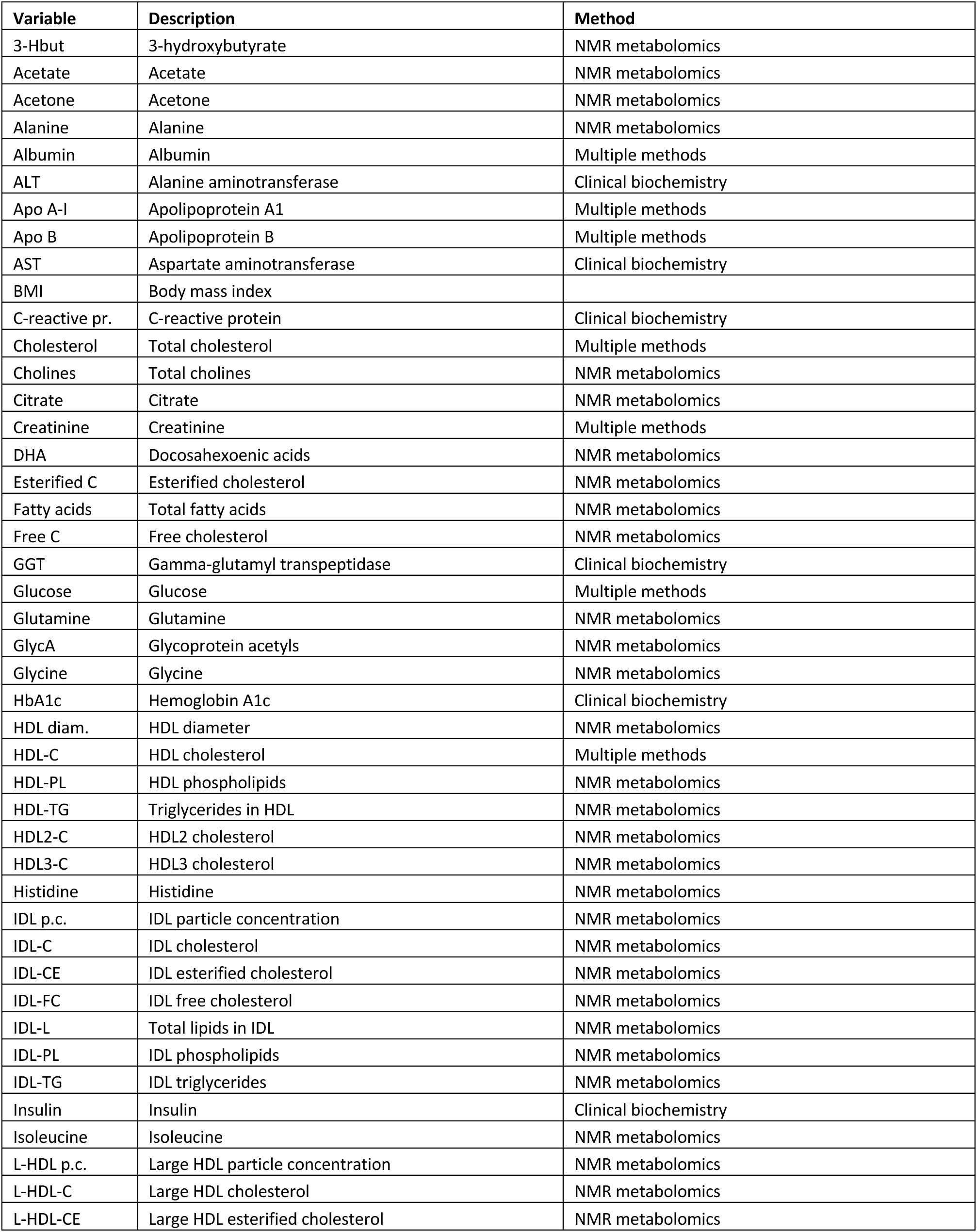
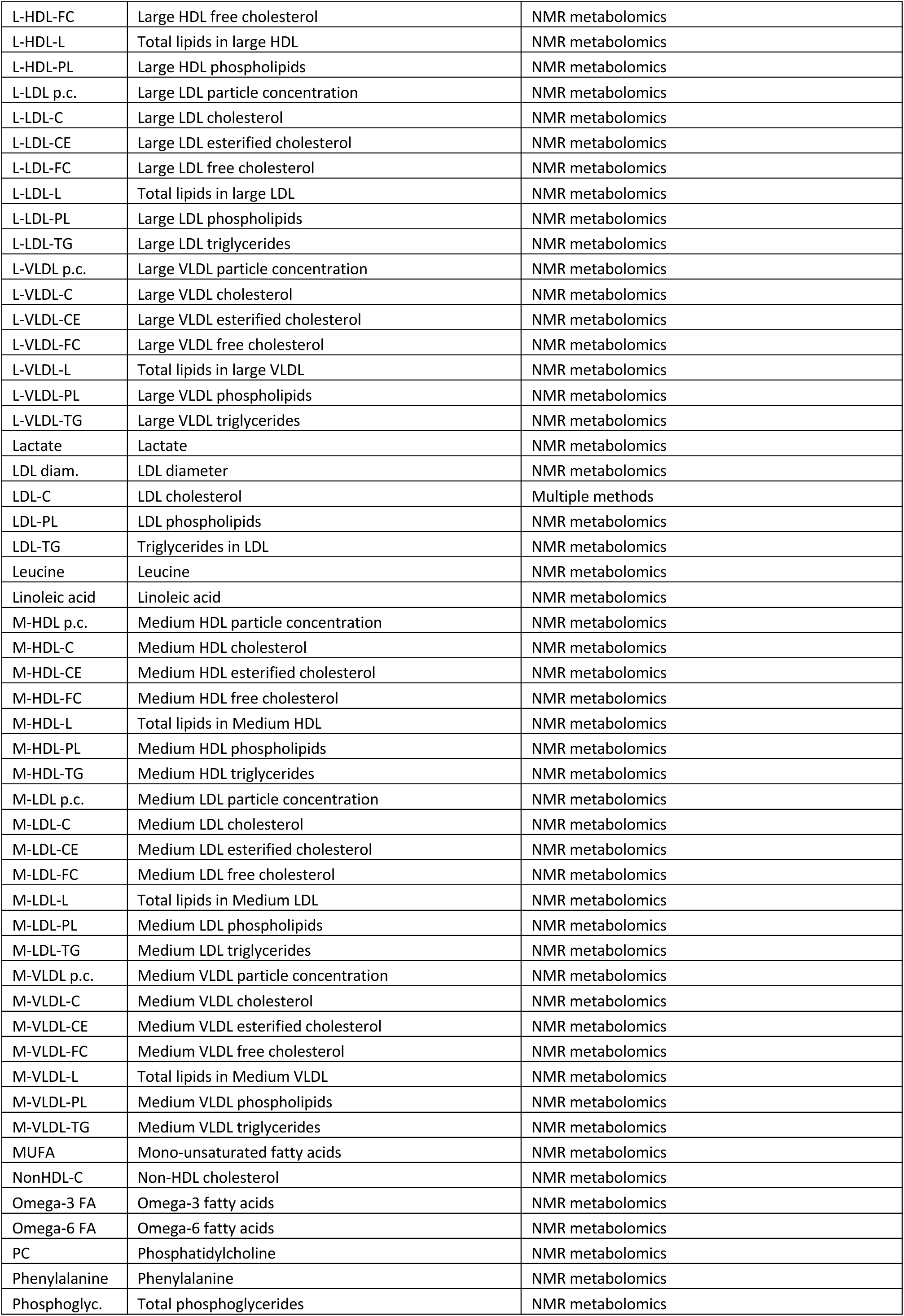
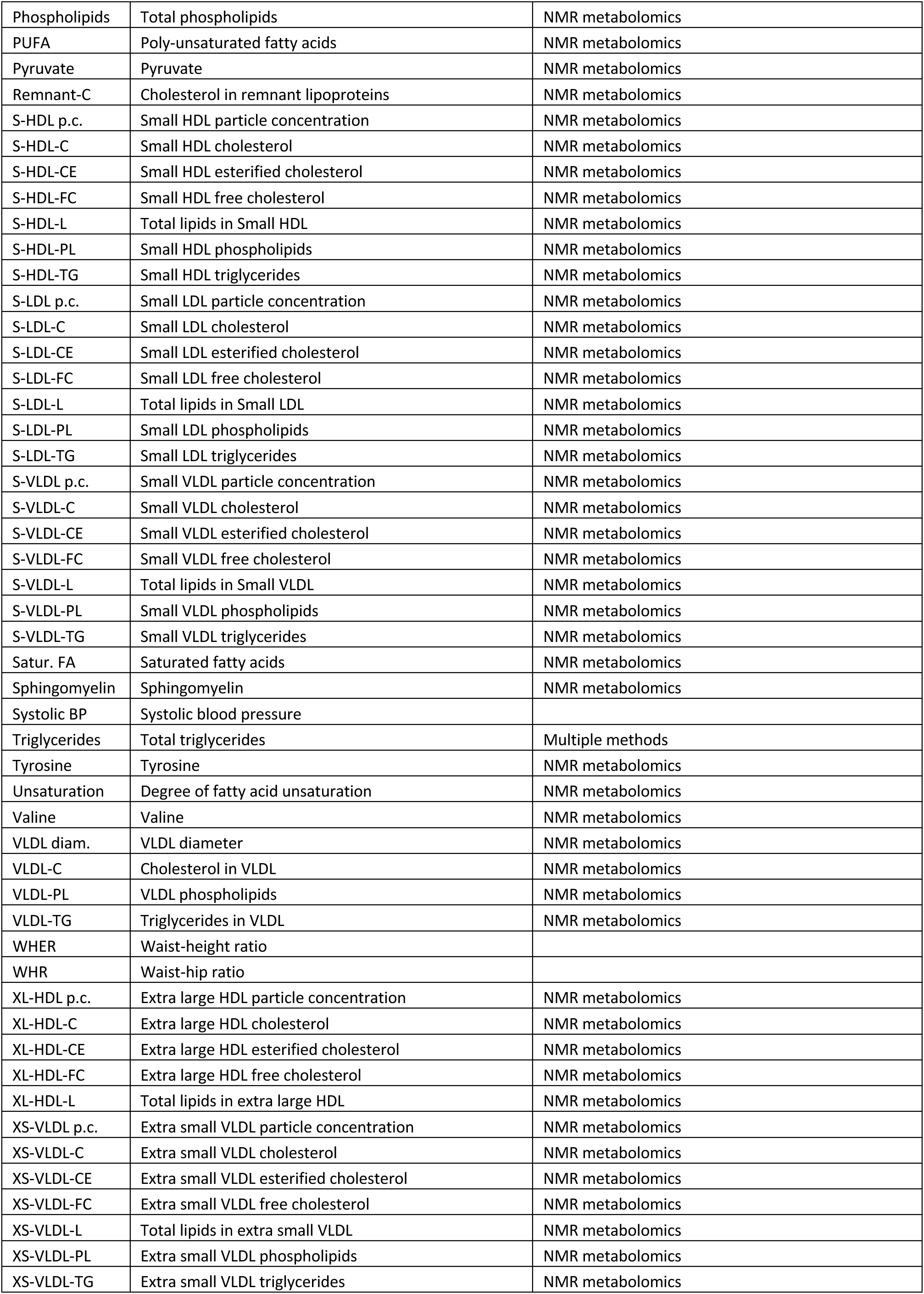
Selection of data for training the temporal self-organizing map. Most measures were available in all cohorts with sufficient coverage across participants. Measures that contained a high frequency of zero concentrations were excluded (e.g. the two largest very-low-density lipoprotein subclasses). Hemoglobin A1c was not available in ALSPAC or YFS cohorts. Liver enzymes (ALT, AST and GGT) were not available in the YFS cohort. Insulin was not available in the UK Biobank.

**Table S3:** Cochrane reviews for anti-hypertensive medications. These consensus effect sizes were the basis for blood pressure adjustments in the UK Biobank.

| Report | Drug type | Systolic effect (mmHg) | Diastolic effect (mmHg) |
| --- | --- | --- | --- |
| Heran BS, Wong MMY, Heran IK, Wright JM. Blood pressure lowering efficacy of angiotensin converting enzyme (ACE) inhibitors for primary hypertension. The Cochrane Library 2009, Issue 4 | Angiotensin-converting enzyme inhibitor | -8 | -5 |
| Heran BS, Wong MMY, Heran IK, Wright JM. Blood pressure lowering efficacy of angiotensin receptor blockers for primary hypertension. | Angiotensin receptor blocker | -10 | -7 |
| Ghamami N, Chiang SHY, Dormuth C, Wright JM. Time course for blood pressure lowering of dihydropyridine calcium channel blockers (Review). The Cochrane Library 2014, Issue 8. The Cochrane Library 2009, Issue 4 | Calcium channel blocker | -10 | -7 |
| Chen JMH, Heran BS, Wright JM. Cochrane Blood pressure lowering efficacy of diuretics as second-line therapy for primary hypertension. Database of Systematic Reviews 2009, Issue4. Art.No.: CD007187 | Diuretics | -7 | -4 |
| Wong GWK, Wright JM. Blood pressure lowering efficacy of nonselective beta-blockers for primary hypertension. Cochrane Database of Systematic Reviews 2014, Issue2 Art.No.: CD007452 | Beta blocker | -10 | -7 |

**Table S4:** Prevalence of clinical classifications within metabolic subgroups (UK Biobank). Diagnosis codes are from the International Statistical Classification of Diseases and Related Health Problems, 10th Revision.

| Subgroup | Diagnosis | Male | Number | Frequency | SE | CI95 low | CI95 high |
| --- | --- | --- | --- | --- | --- | --- | --- |
| High HDL ApoB- | E11 | 1 | 1085 | 0.6% | 0.2% | 0.2% | 1.0% |
| High HDL ApoB- | E11 | 0 | 1239 | 0.2% | 0.1% | 0.0% | 0.4% |
| High HDL ApoB- | E66 | 1 | 1085 | 0.2% | 0.1% | 0.0% | 0.5% |
| High HDL ApoB- | E66 | 0 | 1239 | 0.6% | 0.2% | 0.2% | 1.0% |
| High HDL ApoB- | E78 | 1 | 1085 | 10.1% | 0.9% | 8.4% | 12.0% |
| High HDL ApoB- | E78 | 0 | 1239 | 4.5% | 0.6% | 3.4% | 5.5% |
| High HDL ApoB- | I10 | 1 | 1085 | 17.1% | 1.3% | 14.8% | 19.8% |
| High HDL ApoB- | I10 | 0 | 1239 | 12.1% | 1.0% | 10.4% | 14.0% |
| High HDL ApoB- | I25 | 1 | 1085 | 2.5% | 0.5% | 1.6% | 3.4% |
| High HDL ApoB- | I25 | 0 | 1239 | 0.5% | 0.2% | 0.2% | 0.8% |
| High HDL ApoB+ | E11 | 1 | 847 | 4.1% | 0.7% | 2.7% | 5.5% |
| High HDL ApoB+ | E11 | 0 | 894 | 2.0% | 0.5% | 1.0% | 2.9% |
| High HDL ApoB+ | E66 | 1 | 847 | 1.5% | 0.4% | 0.8% | 2.4% |
| High HDL ApoB+ | E66 | 0 | 894 | 0.2% | 0.2% | 0.0% | 0.7% |
| High HDL ApoB+ | E78 | 1 | 847 | 29.9% | 1.7% | 26.6% | 32.9% |
| High HDL ApoB+ | E78 | 0 | 894 | 16.0% | 1.3% | 13.6% | 18.6% |
| High HDL ApoB+ | I10 | 1 | 847 | 37.4% | 1.6% | 34.1% | 40.6% |
| High HDL ApoB+ | I10 | 0 | 894 | 23.2% | 1.4% | 20.6% | 25.7% |
| High HDL ApoB+ | I25 | 1 | 847 | 4.5% | 0.7% | 3.1% | 5.9% |
| High HDL ApoB+ | I25 | 0 | 894 | 0.8% | 0.3% | 0.2% | 1.5% |
| High LDL | E11 | 1 | 1076 | 0.8% | 0.3% | 0.3% | 1.5% |
| High LDL | E11 | 0 | 957 | 0.8% | 0.3% | 0.3% | 1.5% |
| High LDL | E66 | 1 | 1076 | 0.8% | 0.3% | 0.4% | 1.3% |
| High LDL | E66 | 0 | 957 | 0.4% | 0.2% | 0.1% | 0.8% |
| High LDL | E78 | 1 | 1076 | 43.3% | 1.5% | 40.4% | 46.8% |
| High LDL | E78 | 0 | 957 | 27.6% | 1.4% | 25.2% | 30.4% |
| High LDL | I10 | 1 | 1076 | 35.3% | 1.5% | 32.1% | 37.9% |
| High LDL | I10 | 0 | 957 | 24.1% | 1.2% | 21.8% | 26.0% |
| High LDL | I25 | 1 | 1076 | 4.6% | 0.6% | 3.3% | 6.0% |
| High LDL | I25 | 0 | 957 | 1.2% | 0.4% | 0.5% | 2.0% |
| Low lipid | E11 | 1 | 915 | 1.7% | 0.4% | 1.0% | 2.6% |
| Low lipid | E11 | 0 | 828 | 0.3% | 0.2% | 0.0% | 0.7% |
| Low lipid | E66 | 1 | 915 | 1.3% | 0.4% | 0.5% | 2.1% |
| Low lipid | E66 | 0 | 828 | 0.7% | 0.3% | 0.1% | 1.3% |
| Low lipid | E78 | 1 | 915 | 10.5% | 1.1% | 8.5% | 12.6% |
| Low lipid | E78 | 0 | 828 | 4.1% | 0.7% | 2.8% | 5.4% |
| Low lipid | I10 | 1 | 915 | 26.9% | 1.5% | 24.3% | 30.1% |
| Low lipid | I10 | 0 | 828 | 15.2% | 1.2% | 12.7% | 17.5% |
| Low lipid | I25 | 1 | 915 | 3.5% | 0.6% | 2.5% | 4.7% |
| Low lipid | I25 | 0 | 828 | 0.2% | 0.2% | 0.0% | 0.6% |
| MetDys TG- | E11 | 1 | 1095 | 10.2% | 0.9% | 8.6% | 12.0% |
| MetDys TG- | E11 | 0 | 938 | 5.1% | 0.7% | 3.8% | 6.6% |
| MetDys TG- | E66 | 1 | 1095 | 3.3% | 0.6% | 2.2% | 4.4% |
| MetDys TG- | E66 | 0 | 938 | 5.0% | 0.7% | 3.6% | 6.4% |
| MetDys TG- | E78 | 1 | 1095 | 23.5% | 1.2% | 21.1% | 25.9% |
| MetDys TG- | E78 | 0 | 938 | 13.1% | 1.1% | 11.0% | 14.9% |
| MetDys TG- | I10 | 1 | 1095 | 47.9% | 1.6% | 45.0% | 50.7% |
| MetDys TG- | I10 | 0 | 938 | 37.6% | 1.6% | 35.0% | 40.7% |
| MetDys TG- | I25 | 1 | 1095 | 5.8% | 0.8% | 4.5% | 7.2% |
| MetDys TG- | I25 | 0 | 938 | 1.8% | 0.4% | 1.1% | 2.7% |
| MetDys TG+ | E11 | 1 | 1095 | 7.0% | 0.8% | 5.6% | 8.7% |
| MetDys TG+ | E11 | 0 | 1085 | 4.1% | 0.6% | 2.9% | 5.3% |
| MetDys TG+ | E66 | 1 | 1095 | 2.3% | 0.5% | 1.6% | 3.2% |
| MetDys TG+ | E66 | 0 | 1085 | 3.4% | 0.6% | 2.3% | 4.4% |
| MetDys TG+ | E78 | 1 | 1095 | 36.5% | 1.5% | 33.7% | 39.2% |
| MetDys TG+ | E78 | 0 | 1085 | 25.0% | 1.4% | 22.3% | 27.7% |
| MetDys TG+ | I10 | 1 | 1095 | 43.5% | 1.5% | 40.3% | 46.1% |
| MetDys TG+ | I10 | 0 | 1085 | 38.1% | 1.4% | 35.1% | 40.6% |
| MetDys TG+ | I25 | 1 | 1095 | 6.5% | 0.8% | 4.8% | 7.9% |
| MetDys TG+ | I25 | 0 | 1085 | 2.0% | 0.4% | 1.3% | 2.9% |
| Reference | E11 | 1 | 931 | 0.4% | 0.2% | 0.1% | 0.9% |
| Reference | E11 | 0 | 810 | 0.1% | 0.1% | 0.0% | 0.4% |
| Reference | E66 | 1 | 931 | 0.9% | 0.3% | 0.4% | 1.5% |
| Reference | E66 | 0 | 810 | 0.8% | 0.3% | 0.2% | 1.4% |
| Reference | E78 | 1 | 931 | 15.6% | 1.2% | 13.4% | 17.9% |
| Reference | E78 | 0 | 810 | 6.9% | 0.9% | 5.3% | 8.9% |
| Reference | I10 | 1 | 931 | 25.3% | 1.4% | 22.9% | 28.1% |
| Reference | I10 | 0 | 810 | 15.6% | 1.3% | 13.2% | 18.0% |
| Reference | I25 | 1 | 931 | 3.0% | 0.6% | 2.0% | 4.1% |
| Reference | I25 | 0 | 810 | 0.1% | 0.1% | 0.0% | 0.4% |

**Table S5:** Incidence of clinical classifications within metabolic subgroups (UK Biobank; 14.5 years of follow-up). Diagnosis codes are from the International Statistical Classification of Diseases and Related Health Problems, 10th Revision. Numbers of disease-free individuals at baseline are listed.

| Subgroup | Diagnosis | Male | Number | Frequency | SE | CI95 low | CI95 high |
| --- | --- | --- | --- | --- | --- | --- | --- |
| High HDL ApoB- | E11 | 0 | 1237 | 0.5% | 0.2% | 0.2% | 0.9% |
| High HDL ApoB- | E11 | 1 | 1079 | 1.5% | 0.4% | 0.8% | 2.2% |
| High HDL ApoB- | E66 | 0 | 1232 | 0.5% | 0.2% | 0.2% | 0.9% |
| High HDL ApoB- | E66 | 1 | 1083 | 1.1% | 0.3% | 0.6% | 1.7% |
| High HDL ApoB- | E78 | 0 | 1183 | 7.7% | 0.9% | 6.0% | 9.4% |
| High HDL ApoB- | E78 | 1 | 975 | 13.6% | 1.1% | 11.3% | 15.8% |
| High HDL ApoB- | I10 | 0 | 1089 | 11.2% | 0.9% | 9.4% | 12.9% |
| High HDL ApoB- | I10 | 1 | 899 | 15.7% | 1.2% | 13.7% | 18.0% |
| High HDL ApoB- | I25 | 0 | 1233 | 2.1% | 0.4% | 1.4% | 2.8% |
| High HDL ApoB- | I25 | 1 | 1058 | 6.4% | 0.8% | 4.9% | 7.9% |
| High HDL ApoB+ | E11 | 0 | 876 | 2.4% | 0.5% | 1.5% | 3.5% |
| High HDL ApoB+ | E11 | 1 | 812 | 4.6% | 0.7% | 3.2% | 6.0% |
| High HDL ApoB+ | E66 | 0 | 892 | 2.4% | 0.5% | 1.5% | 3.3% |
| High HDL ApoB+ | E66 | 1 | 834 | 4.1% | 0.7% | 2.9% | 5.5% |
| High HDL ApoB+ | E78 | 0 | 750 | 14.6% | 1.3% | 12.4% | 17.3% |
| High HDL ApoB+ | E78 | 1 | 593 | 21.0% | 1.5% | 18.4% | 24.1% |
| High HDL ApoB+ | I10 | 0 | 686 | 16.1% | 1.4% | 13.5% | 19.0% |
| High HDL ApoB+ | I10 | 1 | 530 | 21.7% | 1.7% | 18.7% | 25.1% |
| High HDL ApoB+ | I25 | 0 | 887 | 4.0% | 0.6% | 2.8% | 5.4% |
| High HDL ApoB+ | I25 | 1 | 808 | 11.2% | 1.1% | 9.0% | 13.4% |
| High LDL | E11 | 0 | 949 | 2.8% | 0.6% | 1.8% | 3.9% |
| High LDL | E11 | 1 | 1067 | 3.9% | 0.6% | 2.8% | 5.1% |
| High LDL | E66 | 0 | 953 | 4.1% | 0.6% | 2.9% | 5.4% |
| High LDL | E66 | 1 | 1067 | 5.7% | 0.7% | 4.5% | 7.1% |
| High LDL | E78 | 0 | 692 | 23.6% | 1.6% | 20.7% | 26.4% |
| High LDL | E78 | 1 | 608 | 34.0% | 1.7% | 30.8% | 37.2% |
| High LDL | I10 | 0 | 726 | 17.2% | 1.4% | 14.6% | 20.0% |
| High LDL | I10 | 1 | 696 | 25.5% | 1.7% | 22.6% | 29.0% |
| High LDL | I25 | 0 | 946 | 5.1% | 0.7% | 3.8% | 6.7% |
| High LDL | I25 | 1 | 1027 | 12.4% | 1.0% | 10.4% | 14.4% |
| Low lipid | E11 | 0 | 825 | 1.0% | 0.3% | 0.4% | 1.7% |
| Low lipid | E11 | 1 | 900 | 2.5% | 0.5% | 1.6% | 3.7% |
| Low lipid | E66 | 0 | 822 | 3.2% | 0.6% | 2.1% | 4.4% |
| Low lipid | E66 | 1 | 903 | 2.6% | 0.5% | 1.7% | 3.7% |
| Low lipid | E78 | 0 | 794 | 5.8% | 0.9% | 4.4% | 7.4% |
| Low lipid | E78 | 1 | 820 | 12.3% | 1.2% | 10.0% | 14.6% |
| Low lipid | I10 | 0 | 702 | 13.1% | 1.2% | 11.0% | 15.7% |
| Low lipid | I10 | 1 | 670 | 19.3% | 1.5% | 16.9% | 22.1% |
| Low lipid | I25 | 0 | 826 | 3.2% | 0.7% | 2.1% | 4.5% |
| Low lipid | I25 | 1 | 882 | 9.5% | 1.0% | 7.8% | 11.3% |
| MetDys TG- | E11 | 0 | 890 | 8.5% | 0.9% | 6.5% | 10.3% |
| MetDys TG- | E11 | 1 | 984 | 14.9% | 1.0% | 13.0% | 17.2% |
| MetDys TG- | E66 | 0 | 892 | 19.1% | 1.3% | 16.5% | 21.4% |
| MetDys TG- | E66 | 1 | 1059 | 16.0% | 1.1% | 13.7% | 17.8% |
| MetDys TG- | E78 | 0 | 814 | 12.8% | 1.2% | 10.6% | 15.1% |
| MetDys TG- | E78 | 1 | 838 | 19.7% | 1.3% | 17.5% | 22.4% |
| MetDys TG- | I10 | 0 | 584 | 24.9% | 1.7% | 21.9% | 28.3% |
| MetDys TG- | I10 | 1 | 571 | 32.3% | 2.0% | 28.4% | 35.7% |
| MetDys TG- | I25 | 0 | 921 | 4.9% | 0.7% | 3.7% | 6.2% |
| MetDys TG- | I25 | 1 | 1032 | 14.8% | 1.2% | 12.8% | 17.2% |
| MetDys TG+ | E11 | 0 | 1041 | 10.6% | 0.9% | 8.8% | 12.4% |
| MetDys TG+ | E11 | 1 | 1019 | 15.0% | 1.2% | 12.5% | 17.1% |
| MetDys TG+ | E66 | 0 | 1048 | 16.3% | 1.2% | 14.0% | 18.4% |
| MetDys TG+ | E66 | 1 | 1069 | 12.6% | 1.0% | 10.7% | 14.5% |
| MetDys TG+ | E78 | 0 | 814 | 24.2% | 1.5% | 21.1% | 27.3% |
| MetDys TG+ | E78 | 1 | 697 | 27.7% | 1.6% | 24.1% | 30.9% |
| MetDys TG+ | I10 | 0 | 670 | 23.6% | 1.6% | 20.4% | 26.4% |
| MetDys TG+ | I10 | 1 | 619 | 29.5% | 1.7% | 26.2% | 33.0% |
| MetDys TG+ | I25 | 0 | 1065 | 6.4% | 0.8% | 4.8% | 7.9% |
| MetDys TG+ | I25 | 1 | 1024 | 12.9% | 1.0% | 10.9% | 15.0% |
| Reference | E11 | 0 | 809 | 0.8% | 0.3% | 0.2% | 1.4% |
| Reference | E11 | 1 | 927 | 2.2% | 0.5% | 1.3% | 3.3% |
| Reference | E66 | 0 | 804 | 2.7% | 0.6% | 1.6% | 3.7% |
| Reference | E66 | 1 | 922 | 2.7% | 0.6% | 1.7% | 3.9% |
| Reference | E78 | 0 | 754 | 10.7% | 1.2% | 8.6% | 13.3% |
| Reference | E78 | 1 | 787 | 21.1% | 1.4% | 18.4% | 23.6% |
| Reference | I10 | 0 | 683 | 10.3% | 1.2% | 8.2% | 12.9% |
| Reference | I10 | 1 | 697 | 16.4% | 1.4% | 13.5% | 19.2% |
| Reference | I25 | 0 | 809 | 2.4% | 0.6% | 1.5% | 3.7% |
| Reference | I25 | 1 | 903 | 9.0% | 1.0% | 7.2% | 10.7% |

